# Spatially localized racial differences in the association between white matter hyperintensities and verbal memory in cognitively unimpaired adults

**DOI:** 10.64898/2026.11.10.26367467

**Authors:** Junjie Wu, Qixiang Lin, Taylor A James, Allan I Levey, James J Lah, Deqiang Qiu, The HABS-HD Study Team

## Abstract

White matter hyperintensities (WMH) are common in aging, but whether their cognitive associations differ between African American and White older adults is unclear. We studied 5,187 cognitively unimpaired non-Hispanic African American (NHA) and non-Hispanic White (NHW) older adults across three independent cohorts using depth- and lobe-resolved WMH mapping with cross-sectional, longitudinal and machine learning analyses. In the Emory Healthy Brain Study (EHBS), greater WMH burden was more strongly associated with poorer verbal memory among NHA than NHW participants. A similar difference was observed in a 34-center cohort but not detected in the third cohort. Differences localized to frontal juxtacortical and parietal deep white matter in EHBS and the multicenter cohort, respectively, and did not parallel WMH burden distributions. Estimates changed little after adjustment for cohort-specific Alzheimer’s disease (AD) biomarkers, vascular risk factors, structural imaging measures and *APOE* genotype. Accounting for neighborhood deprivation in EHBS yielded a similar estimate. Generalized random forests supported these differences without assuming a linear interaction. Baseline frontal juxtacortical WMH burden showed a stronger association with subsequent AD-signature cortical thinning among NHA than NHW participants in EHBS, with directionally similar multicenter findings. Spatially resolved WMH measures may help identify early heterogeneity in vascular contributions to cognitive aging.

## 1. Introduction

White matter hyperintensities (WMH) are a common neuroimaging marker of cerebral small vessel disease in later life^1,2^ and are associated with cognitive decline, stroke, and incident dementia^2^. In relatively healthy older adults, greater WMH burden may precede declines in memory, attention, and executive function^3^. Examining the association between WMH burden and cognition before the onset of mild cognitive impairment or dementia would therefore help characterize early differences in the vascular contributions to cognitive aging.

Whether this association differs between African American and White older adults remains unclear. Previous studies have reported stronger associations between WMH burden and poorer performance in language, processing speed or executive function among African American than White participants^4^, as well as faster decline across several cognitive domains among African Americans^5^. In contrast, studies using direct interaction tests^6^ or a white matter connectivity measure^7^ found no evidence of racial differences. Longitudinal studies further indicate that the contribution of WMH to cognitive decline varies across racial and ethnic groups^8^ and could be modified by factors such as education^9^. Variation in study populations, cognitive measures, and analytical approaches has made it difficult to establish whether racial differences in the association between WMH and cognition are reproducible across independent cohorts.

Most previous studies have characterized WMH using total or global burden, which may obscure clinically relevant spatial variation. Evidence increasingly suggests that WMH location is biologically informative, with different spatial distributions associated with vascular, amyloid, and neurodegenerative processes^10–13^. WMH location also appears relevant to cognition, as lesions in different white matter regions show distinct associations with cognitive performance and decline even after accounting for total WMH burden^13–15^. Spatially resolved analyses can therefore clarify whether racial differences in the association between WMH burden and cognition vary across white matter locations and whether similar spatial patterns are observed across cohorts.

In this study, we examined cognitively unimpaired non-Hispanic African American (NHA) and non-Hispanic White (NHW) older adults across three large independent cohorts to determine whether the association between WMH and cognition differed by race and whether these differences were localized to specific white matter depths and lobes. We then assessed whether the observed differences persisted after accounting for Alzheimer’s disease (AD) biomarkers, structural imaging measures, apolipoprotein E (*APOE*) genotype, vascular risk factors and neighborhood deprivation. Machine learning analyses further evaluated the robustness of the findings beyond linear models. Finally, longitudinal analyses examined whether baseline WMH burden was associated with subsequent structural change and whether this association also differed by race.

## 2. Results

### 2.1 Study overview and cohort characteristics

The study included cognitively unimpaired NHA and NHW participants from the Emory Healthy Brain Study (EHBS), National Alzheimer’s Coordinating Center (NACC) and Health and Aging Brain Study–Health Disparities (HABS-HD) (Fig. 1a). WMH were segmented and localized by lobe and white matter depth using a four-lobe, four-layer parcellation (Fig. 1b,c). Cross-sectional analyses examined racial differences in the associations between WMH burden and neuropsychological performance. Longitudinal analyses examined racial differences in the associations between baseline WMH burden and subsequent changes in matched-lobe cortical thickness, AD-signature cortical thickness (Fig. 1d) and hippocampal volume.

**Fig. 1.**
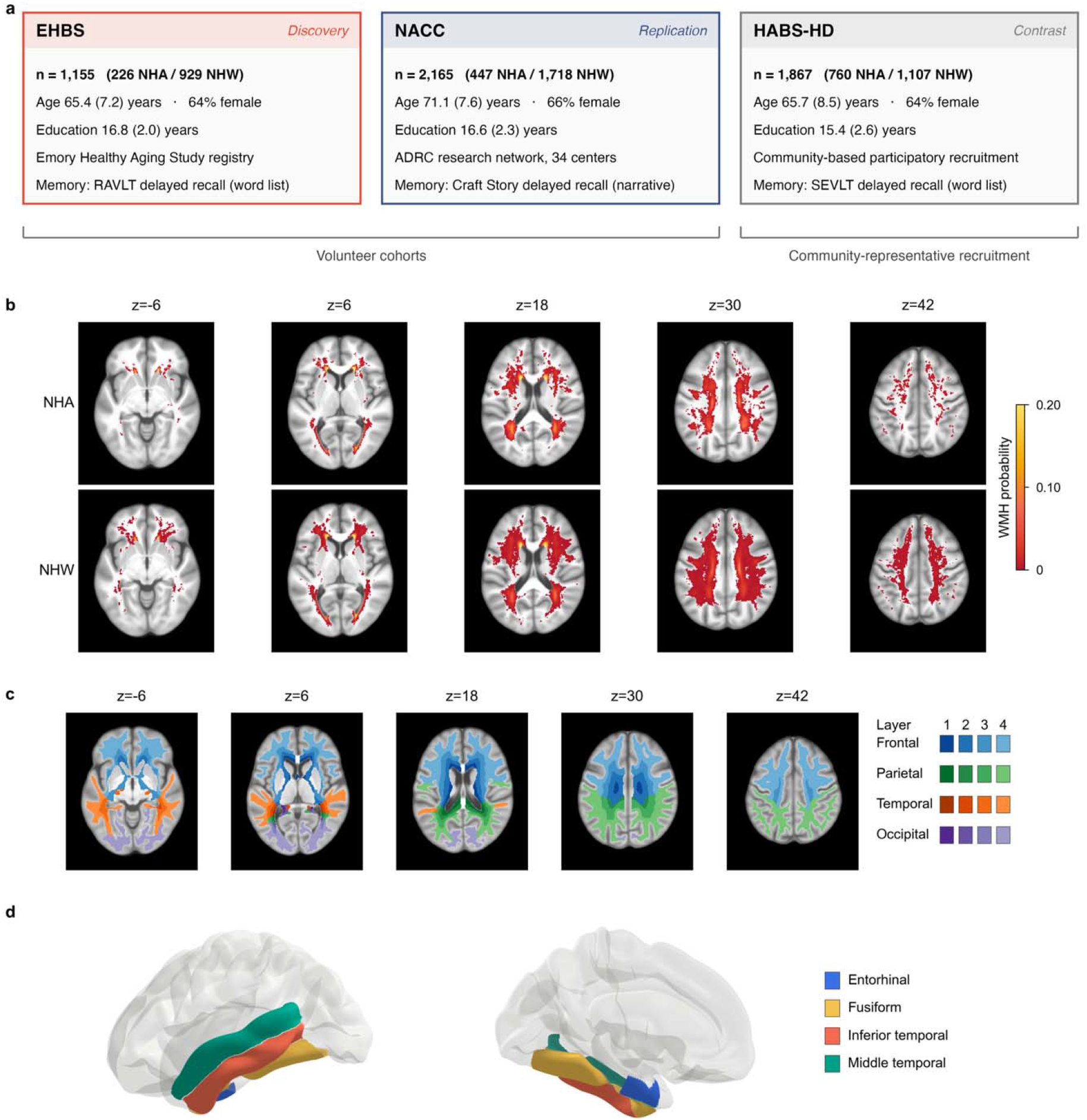
Study design and imaging framework. **a**, Overview of the three study cohorts. Values are mean (s.d.) or percentage. **b**, Race-stratified WMH probability maps in EHBS. **c**, Four-lobe, four-layer white matter parcellation used to localize WMH, with layer 1 representing periventricular, layers 2–3 deep and layer 4 juxtacortical white matter. **d**, Entorhinal, fusiform, inferior temporal and middle temporal regions defining the AD-signature cortex. Panels **b**–**d** use EHBS anatomy for illustration. The same WMH segmentation and parcellation procedures were applied across cohorts. EHBS, Emory Healthy Brain Study; HABS-HD, Health and Aging Brain Study–Health Disparities; NACC, National Alzheimer’s Coordinating Center; NHA, non-Hispanic African American; NHW, non-Hispanic White; RAVLT, Rey Auditory Verbal Learning Test; SEVLT, Spanish-English Verbal Learning Test; WMH, white matter hyperintensities.

The analytic sample comprised 5,187 participants, including 1,155 from EHBS (226 NHA and 929 NHW), 2,165 from NACC (447 NHA and 1,718 NHW) across 34 Alzheimer’s Disease Research Centers, and 1,867 from HABS-HD (760 NHA and 1,107 NHW) (Fig. 1a and Supplementary Fig. 1). Across all three cohorts, NHA participants had fewer years of education and higher prevalences of hypertension, diabetes and obesity compared to NHW participants, as well as poorer performance across cognitive domains (Table 1 and Supplementary Tables 1 and 2). Neighborhood deprivation, assessed using the Area Deprivation Index (ADI), was also higher among NHA than NHW participants.

**Table 1.**
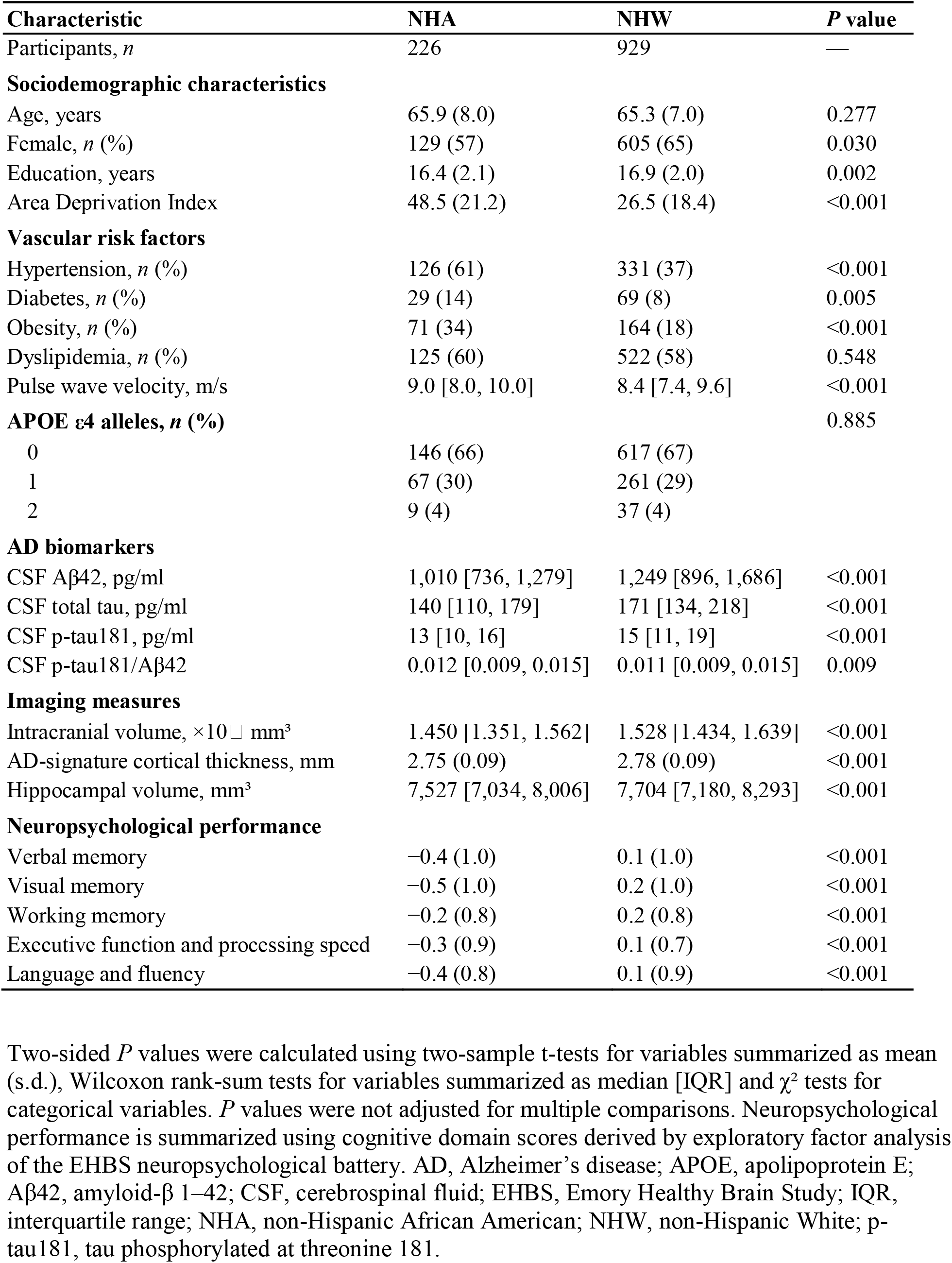
Participant characteristics in EHBS.

| Characteristic | NHA | NHW | *P* value |
|---|---|---|---|
| Participants, *n* | 226 | 929 | — |
| **Sociodemographic characteristics** |  |  |  |
| Age, years | 65.9 (8.0) | 65.3 (7.0) | 0.277 |
| Female, *n* (%) | 129 (57) | 605 (65) | 0.030 |
| Education, years | 16.4 (2.1) | 16.9 (2.0) | 0.002 |
| Area Deprivation Index | 48.5 (21.2) | 26.5 (18.4) | <0.001 |
| **Vascular risk factors** |  |  |  |
| Hypertension, *n* (%) | 126 (61) | 331 (37) | <0.001 |
| Diabetes, *n* (%) | 29 (14) | 69 (8) | 0.005 |
| Obesity, *n* (%) | 71 (34) | 164 (18) | <0.001 |
| Dyslipidemia, *n* (%) | 125 (60) | 522 (58) | 0.548 |
| Pulse wave velocity, m/s | 9.0 [8.0, 10.0] | 8.4 [7.4, 9.6] | <0.001 |
| **APOE ε4 alleles, *n* (%)** |  |  | 0.885 |
| 0 | 146 (66) | 617 (67) |  |
| 1 | 67 (30) | 261 (29) |  |
| 2 | 9 (4) | 37 (4) |  |
| **AD biomarkers** |  |  |  |
| CSF Aβ42, pg/ml | 1,010 [736, 1,279] | 1,249 [896, 1,686] | <0.001 |
| CSF total tau, pg/ml | 140 [110, 179] | 171 [134, 218] | <0.001 |
| CSF p-tau181, pg/ml | 13 [10, 16] | 15 [11, 19] | <0.001 |
| CSF p-tau181/Aβ42 | 0.012 [0.009, 0.015] | 0.011 [0.009, 0.015] | 0.009 |
| **Imaging measures** |  |  |  |
| Intracranial volume, ×10□ mm³ | 1.450 [1.351, 1.562] | 1.528 [1.434, 1.639] | <0.001 |
| AD-signature cortical thickness, mm | 2.75 (0.09) | 2.78 (0.09) | <0.001 |
| Hippocampal volume, mm³ | 7,527 [7,034, 8,006] | 7,704 [7,180, 8,293] | <0.001 |
| **Neuropsychological performance** |  |  |  |
| Verbal memory | −0.4 (1.0) | 0.1 (1.0) | <0.001 |
| Visual memory | −0.5 (1.0) | 0.2 (1.0) | <0.001 |
| Working memory | −0.2 (0.8) | 0.2 (0.8) | <0.001 |
| Executive function and processing speed | −0.3 (0.9) | 0.1 (0.7) | <0.001 |
| Language and fluency | −0.4 (0.8) | 0.1 (0.9) | <0.001 |

Two-sided *P* values were calculated using two-sample t-tests for variables summarized as mean (s.d.), Wilcoxon rank-sum tests for variables summarized as median [IQR] and χ² tests for categorical variables. *P* values were not adjusted for multiple comparisons. Neuropsychological performance is summarized using cognitive domain scores derived by exploratory factor analysis of the EHBS neuropsychological battery. AD, Alzheimer’s disease; APOE, apolipoprotein E; Aβ42, amyloid-β 1–42; CSF, cerebrospinal fluid; EHBS, Emory Healthy Brain Study; IQR, interquartile range; NHA, non-Hispanic African American; NHW, non-Hispanic White; p-tau181, tau phosphorylated at threonine 181.

### 2.2 Racial differences in WMH associations with verbal memory

In EHBS, greater WMH burden in deep white matter (layers 2 and 3) and juxtacortical white matter (layer 4) was more strongly associated with poorer delayed recall on the Rey Auditory Verbal Learning Test (RAVLT) among NHA than NHW participants (layer 2: β = −0.14, 95% confidence intervals (CI) −0.26 to −0.02, false discovery rate (FDR)-adjusted *P* = 0.025; layer 3: β = −0.17, 95% CI −0.28 to −0.05, FDR-adjusted *P* = 0.010; layer 4: β = −0.22, 95% CI −0.35 to −0.09, FDR-adjusted *P* = 0.005) (Fig. 2a). Race-stratified analyses showed corresponding associations between greater WMH burden and poorer delayed recall among NHA participants but not NHW participants (Fig. 2b).

**Fig. 2.**
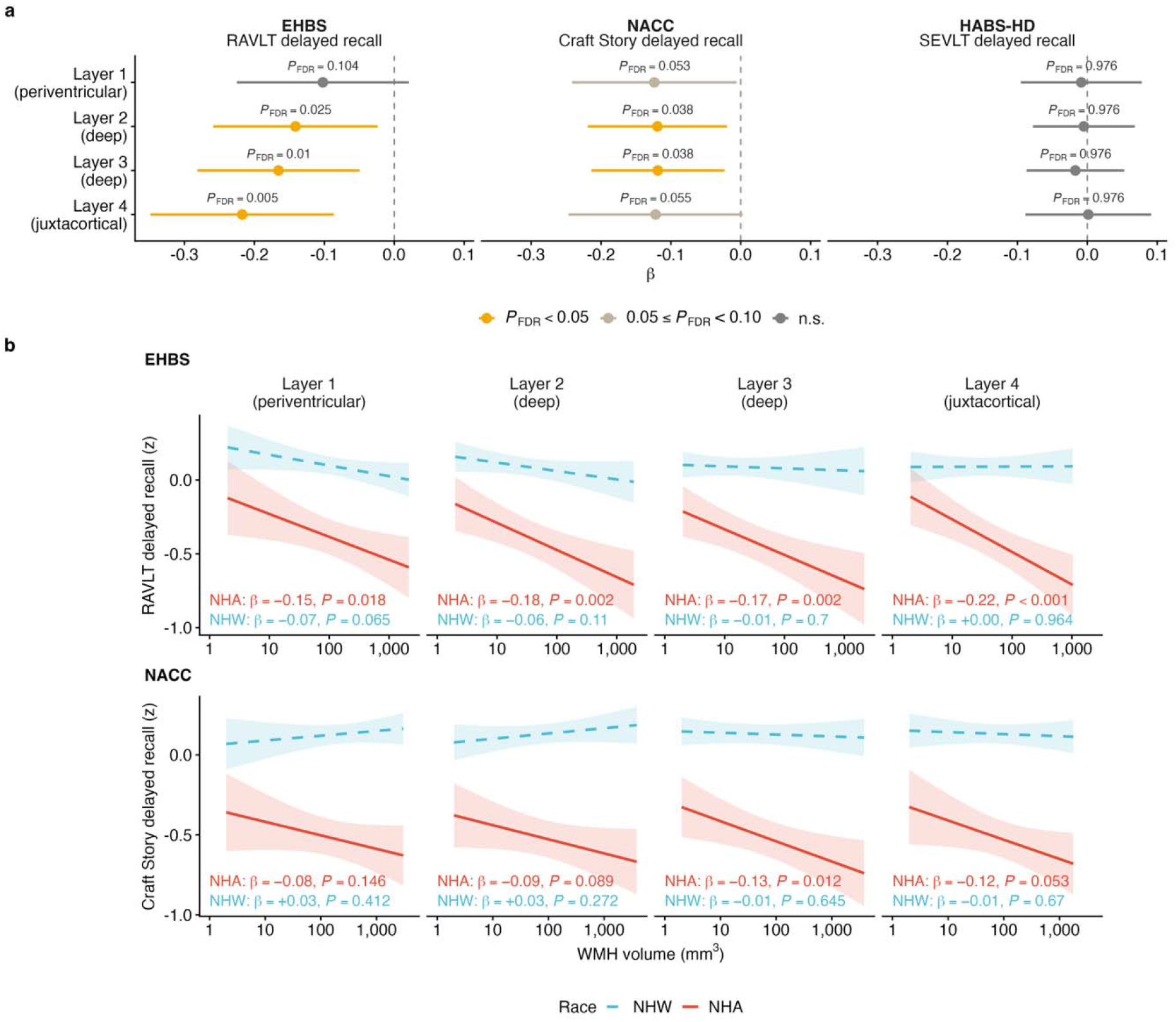
Racial differences in WMH associations with verbal memory across white matter depth. Delayed verbal recall was assessed using RAVLT delayed recall in EHBS, Craft Story delayed recall in NACC and SEVLT delayed recall in HABS-HD. **a**, Interaction between race and WMH burden across four white matter depth layers. Points and bars show interaction coefficients and 95% confidence intervals. Negative coefficients indicate a more negative association among NHA than NHW participants. *P* values were adjusted for false discovery rate. **b**, Race-stratified associations between WMH burden and delayed verbal recall; bands show 95% confidence intervals for predicted means. Linear regression models were adjusted for age, sex, years of education and intracranial volume, with scanner manufacturer additionally included as a fixed effect in NACC. Heteroskedasticity-consistent (HC3) standard errors were used. EHBS, Emory Healthy Brain Study; HABS-HD, Health and Aging Brain Study–Health Disparities; NACC, National Alzheimer’s Coordinating Center; NHA, non-Hispanic African American; NHW, non-Hispanic White; RAVLT, Rey Auditory Verbal Learning Test; SEVLT, Spanish-English Verbal Learning Test; WMH, white matter hyperintensities.

NACC independently showed similar racial differences in the association between deep WMH burden (layers 2 and 3) and Craft Story 21 delayed recall (layer 2: β = −0.12, 95% CI −0.22 to −0.02, FDR-adjusted *P* = 0.038; layer 3: β = −0.12, 95% CI −0.21 to −0.02, FDR-adjusted *P* = 0.038) (Fig. 2a). Race-stratified analyses similarly showed an association between greater layer 3 WMH burden and poorer delayed recall among NHA participants but not NHW participants (Fig. 2b). No corresponding racial difference was detected in HABS-HD.

Of the five cognitive domains examined, only verbal memory showed significant racial differences in both EHBS and NACC after FDR correction (Fig. 3). In EHBS, the interaction between race and WMH burden was significant in layer 4 (β = −0.18, 95% CI −0.31 to −0.05, FDR-adjusted *P* = 0.032). In NACC, significant interactions were observed in layers 2 and 3 (layer 2: β = −0.12, 95% CI −0.22 to −0.02, FDR-adjusted *P* = 0.038; layer 3: β = −0.12, 95% CI −0.21 to −0.02, FDR-adjusted *P* = 0.038). In EHBS, positive interactions for language and fluency were also significant in layers 1 and 2.

**Fig. 3.**
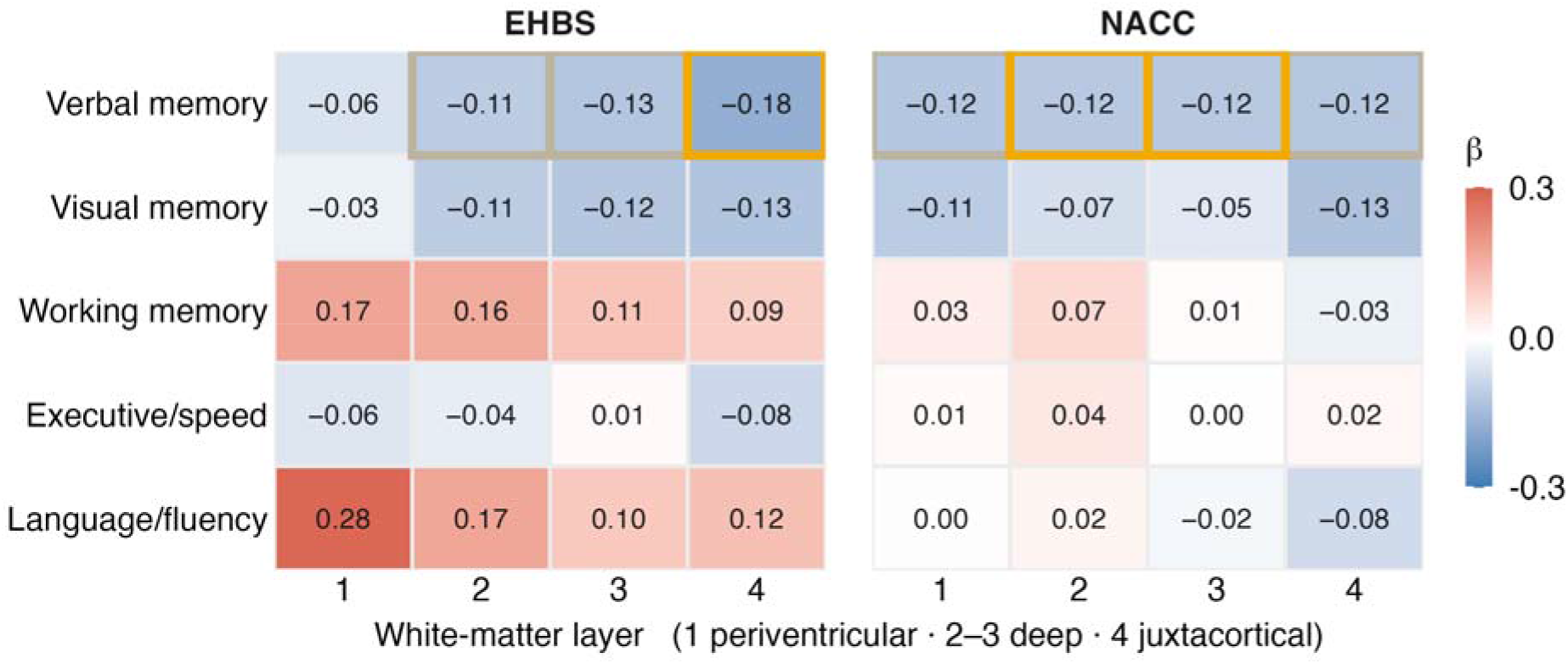
Racial differences in WMH associations across cognitive domains. Interaction coefficients between race and WMH burden are shown across four white matter depth layers and five cognitive domains. Negative coefficients indicate a more negative association among NHA than NHW participants. Orange and grey outlines indicate negative coefficients with *P* values < 0.05 and 0.05–0.10, respectively, after false discovery rate correction. In EHBS, cognitive domain scores representing verbal memory, visual memory, working memory, executive function and processing speed, and language and fluency were derived using exploratory factor analysis. In NACC, Craft Story delayed recall, Benson Complex Figure delayed recall and Number Span Backward represented verbal memory, visual memory and working memory, respectively; harmonized executive function and language scores from the Alzheimer’s Disease Sequencing Project Phenotype Harmonization Consortium were used for the corresponding domains. Linear regression models were adjusted for age, sex, years of education and intracranial volume, with scanner manufacturer additionally included as a fixed effect in NACC. Heteroskedasticity-consistent (HC3) standard errors were used. EHBS, Emory Healthy Brain Study; NACC, National Alzheimer’s Coordinating Center; WMH, white matter hyperintensities.

Lobar analyses further localized these depth-specific differences (Fig. 4a). In EHBS, the racial difference localized to frontal juxtacortical white matter (layer 4) (β = −0.19, 95% CI −0.32 to −0.06, FDR-adjusted *P* = 0.019). A temporal juxtacortical difference also remained significant after FDR correction, but WMH were present in only 11% of participants in that compartment. In NACC, the racial difference localized to parietal deep white matter (layer 3) (β = −0.12, 95% CI −0.22 to −0.03, FDR-adjusted *P* = 0.039). Within the localized compartments, greater WMH burden was associated with poorer delayed recall among NHA participants (EHBS: β = −0.20, 95% CI −0.31 to −0.08, *P* < 0.001; NACC: β = −0.12, 95% CI −0.21 to −0.03, *P* = 0.013) but not NHW participants (Fig. 4b).

**Fig. 4.**
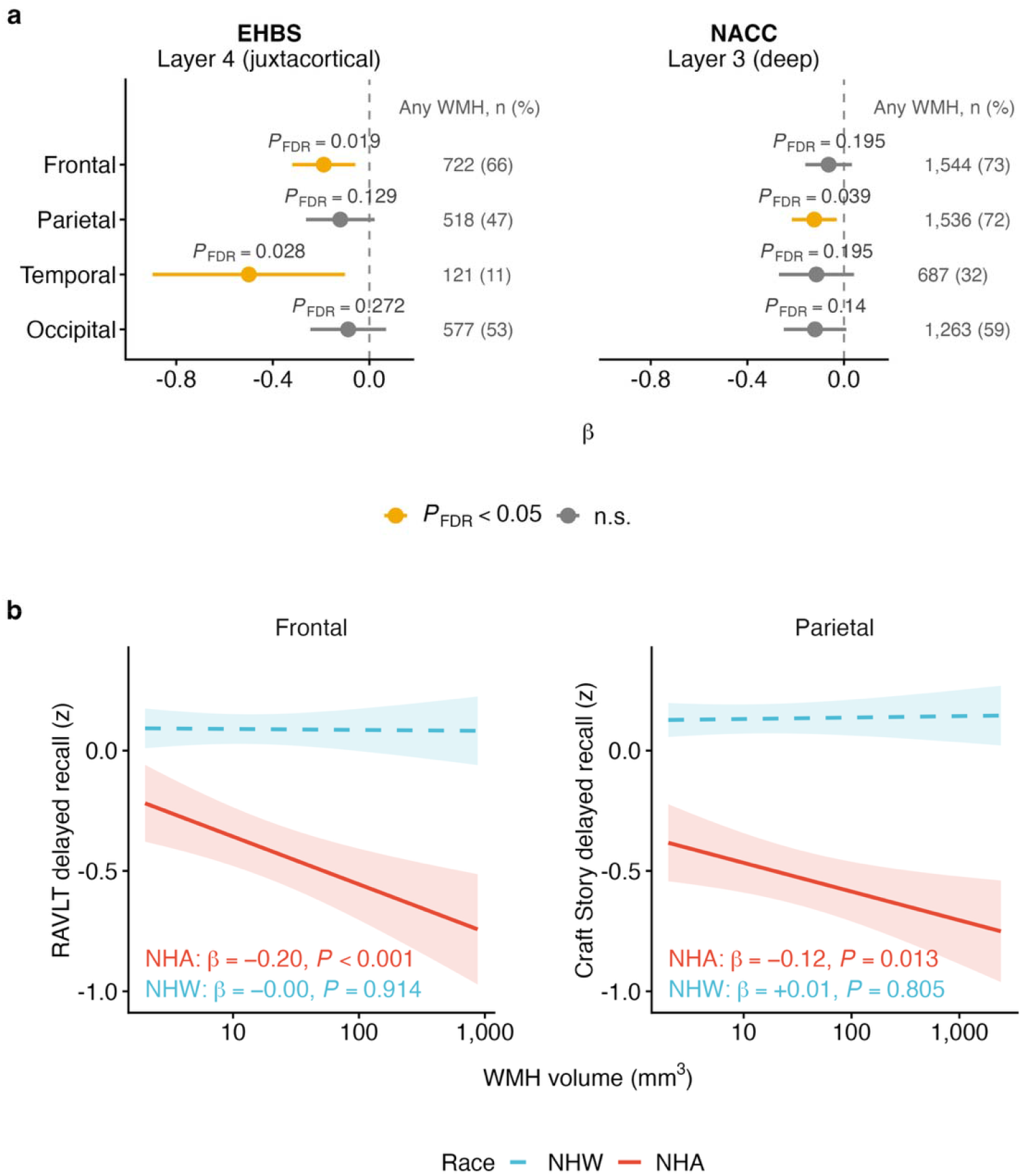
Lobar localization of racial differences in WMH associations with verbal memory. Delayed verbal recall was assessed using RAVLT delayed recall in EHBS and Craft Story delayed recall in NACC. **a**, Interaction between race and WMH burden across four lobes for the depth layers identified in each cohort, juxtacortical white matter (layer 4) in EHBS and deep white matter (layer 3) in NACC. Points and bars show interaction coefficients and 95% confidence intervals. Negative coefficients indicate a more negative association among NHA than NHW participants. *P* values were adjusted for false discovery rate. **b**, Race-stratified associations in frontal juxtacortical white matter (layer 4) in EHBS and parietal deep white matter (layer 3) in NACC; bands show 95% confidence intervals for predicted means. The temporal juxtacortical compartment (layer 4) in EHBS is not shown in **b** because WMH were present in only 11% of participants. Linear regression models were adjusted for age, sex, years of education and intracranial volume, with scanner manufacturer additionally included as a fixed effect in NACC. Heteroskedasticity-consistent (HC3) standard errors were used. EHBS, Emory Healthy Brain Study; NACC, National Alzheimer’s Coordinating Center; NHA, non-Hispanic African American; NHW, non-Hispanic White; RAVLT, Rey Auditory Verbal Learning Test; WMH, white matter hyperintensities.

The localized racial differences did not parallel the spatial distribution of WMH burden. Higher WMH burden among NHA than NHW participants was widespread across white matter regions, particularly in frontal and parietal deep and juxtacortical compartments, across all three cohorts (Supplementary Fig. 2).

### 2.3 Robustness across analytic specifications

In frontal juxtacortical white matter (layer 4) in EHBS and parietal deep white matter (layer 3) in NACC, the racial difference in the association between WMH burden and delayed recall changed little across models additionally adjusted for vascular risk factors (hypertension, diabetes, obesity and dyslipidemia), a cohort-specific AD biomarker, AD-signature cortical thickness, hippocampal volume or *APOE* ε4 (Fig. 5a). The AD biomarker was cerebrospinal fluid (CSF) p-tau181/Aβ42 in EHBS and amyloid positron emission tomography (PET) burden on the Centiloid scale in NACC. In EHBS, adding pulse wave velocity to the vascular risk factors yielded a similar estimate. Adjustment for ADI also yielded a similar estimate, although the interaction was no longer significant after FDR correction; ADI data were missing for 23.0% of NHA and 13.8% of NHW participants. In NACC, estimates remained similar after adjustment for the AD biomarker alone or together with vascular risk factors, although neither analysis remained significant after FDR correction in substantially smaller complete-case samples. Adjustment for *APOE* ε4 likewise yielded a similar estimate but was no longer significant after FDR correction; *APOE* ε4 data were missing for 17.4% of NHA and 4.7% of NHW participants (Supplementary Table 3).

**Fig. 5.**
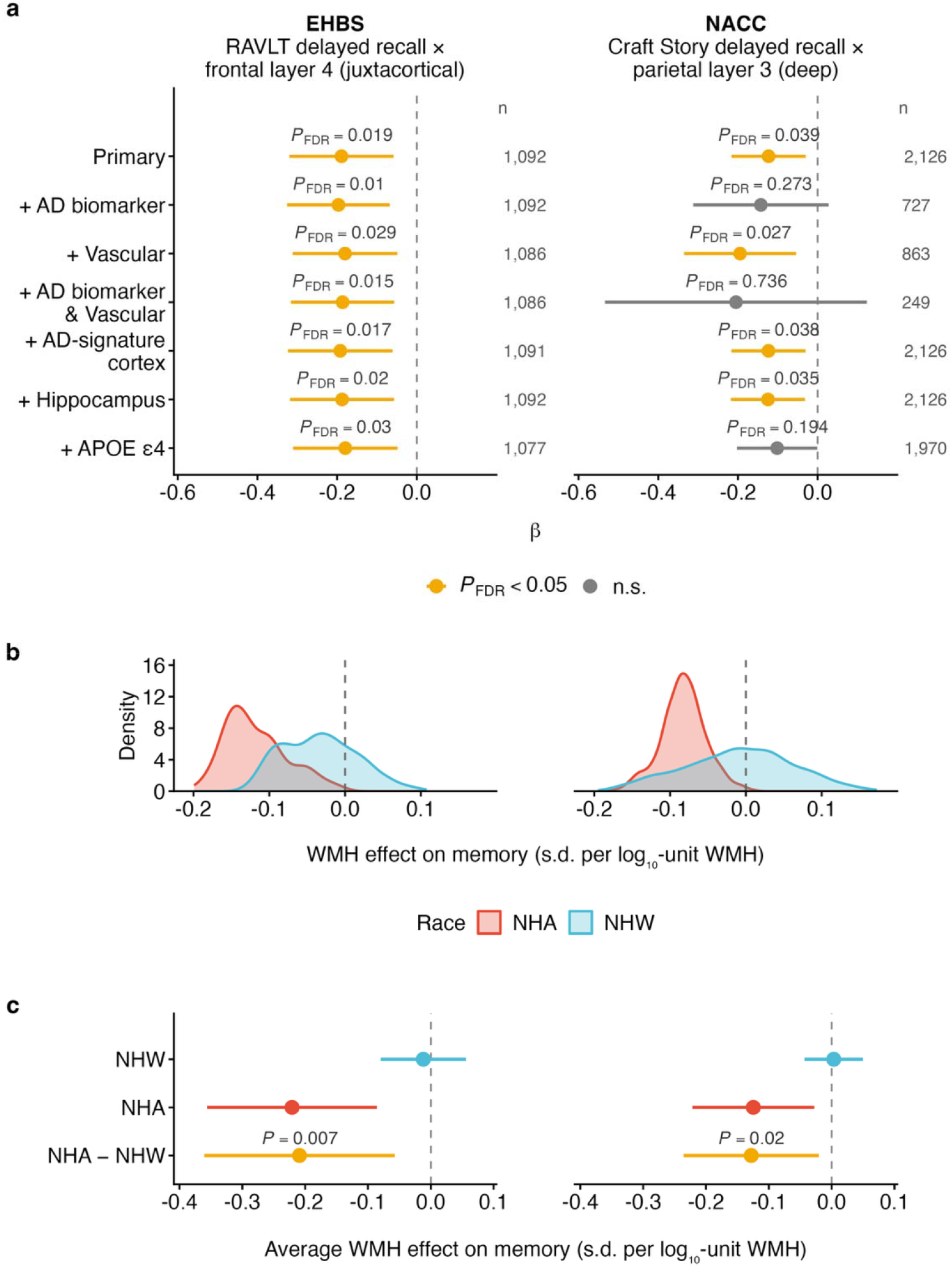
Robustness of racial differences to covariate adjustment and flexible modelling. All panels use frontal juxtacortical white matter (layer 4) in EHBS and parietal deep white matter (layer 3) in NACC. Delayed verbal recall was assessed using RAVLT delayed recall in EHBS and Craft Story delayed recall in NACC. **a**, Racial differences in the association between WMH burden and delayed recall after additional adjustment for vascular risk factors, a cohort-specific AD biomarker (CSF p-tau181/Aβ42 in EHBS and amyloid PET Centiloid in NACC), their combination, AD-signature cortical thickness, hippocampal volume or *APOE* ε4. Points and bars show interaction coefficients and 95% confidence intervals. Negative coefficients indicate a more negative association among NHA than NHW participants. The primary linear regression models were adjusted for age, sex, years of education and intracranial volume, with scanner manufacturer additionally included as a fixed effect in NACC. Heteroskedasticity-consistent (HC3) standard errors were used. *P* values were adjusted for false discovery rate. **b**, Distributions of participant-level conditional effects of WMH burden on delayed recall estimated using generalized random forests, stratified by race. **c**, Doubly robust average partial effects of WMH burden on delayed recall for NHA and NHW participants. The between-group difference was estimated using a best linear projection of the participant-level effects onto race; points and bars show estimates and 95% confidence intervals. AD, Alzheimer’s disease; *APOE*, apolipoprotein E; CSF, cerebrospinal fluid; EHBS, Emory Healthy Brain Study; NACC, National Alzheimer’s Coordinating Center; NHA, non-Hispanic African American; NHW, non-Hispanic White; RAVLT, Rey Auditory Verbal Learning Test; WMH, white matter hyperintensities.

Because NACC included data from 34 centers, we examined sensitivity to alternative specifications for acquisition and center effects. The estimated racial difference remained negative across all specifications (Supplementary Fig. 3a). The center-clustered specification yielded the widest confidence interval; the interaction coefficient remained β = −0.12 (95% CI −0.25 to 0.00) and was no longer significant after FDR correction. When each center was excluded in turn, all interaction coefficients remained negative and of similar magnitude, with the least negative estimate at β = −0.07 (Supplementary Fig. 3b).

Generalized random forests, which allowed the association between WMH burden and delayed recall to vary flexibly without imposing the same linear interaction form, yielded similar racial differences (Fig. 5b,c). The average conditional effect of WMH burden on delayed recall was more negative among NHA than NHW participants, with differences of −0.21 s.d. (95% CI −0.36 to −0.06, *P* = 0.007) in EHBS and −0.13 s.d. (95% CI −0.24 to −0.02, *P* = 0.020) in NACC (Fig. 5c).

### 2.4 Racial differences in WMH associations with subsequent cortical thinning

Longitudinal analyses included 311 EHBS participants (46 NHA) and 518 NACC participants (90 NHA). In EHBS, greater baseline frontal juxtacortical WMH burden (layer 4) was more strongly associated with faster subsequent thinning of the AD-signature cortex among NHA than NHW participants (β = −1.88 × 10^−3^ per year, 95% CI −2.94 × 10^−3^ to −0.81 × 10^−3^, FDR-adjusted *P* = 0.002) (Fig. 6a). Race-stratified analyses showed that greater baseline WMH burden was associated with faster AD-signature cortical thinning among NHA participants (β = −0.38% per year, 95% CI −0.59% to −0.18%, *P* < 0.001) but not NHW participants (Fig. 6b).

**Fig. 6.**
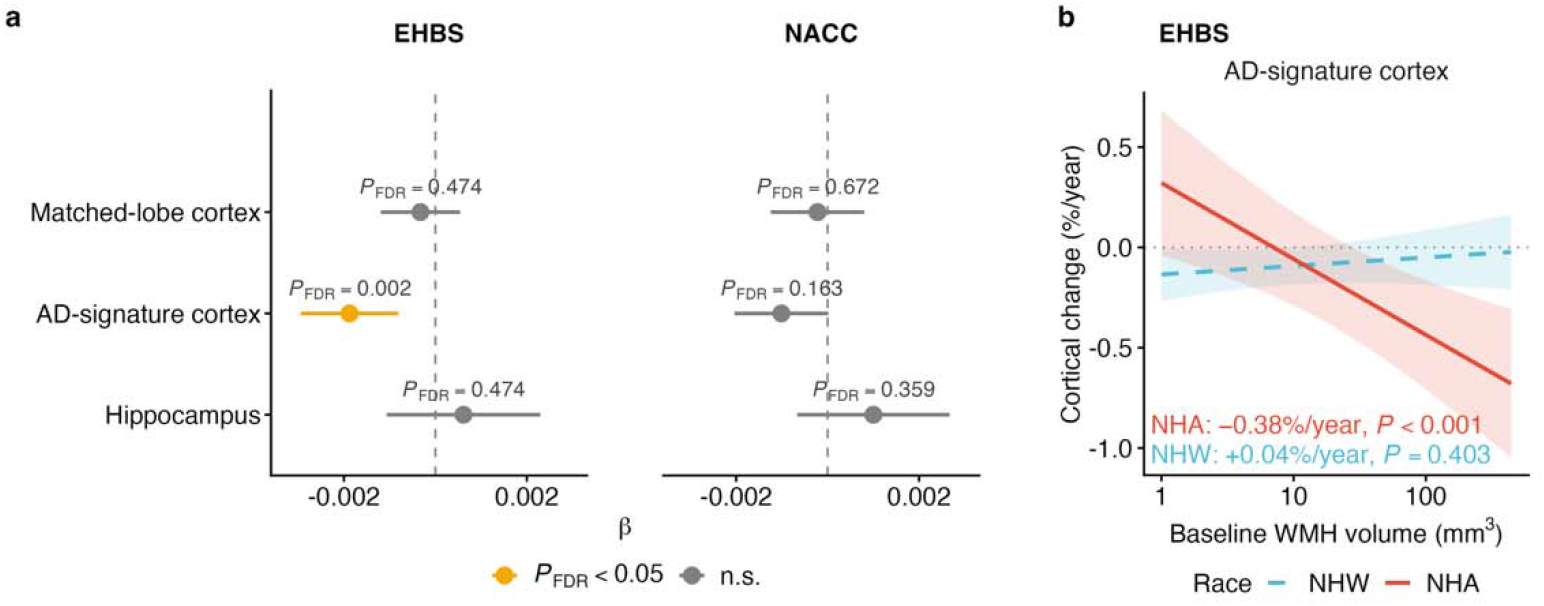
Racial differences in associations of baseline WMH burden with subsequent structural change. **a**, Racial differences in the association of baseline frontal juxtacortical WMH burden (layer 4) with longitudinal change in frontal cortical thickness, AD-signature cortical thickness and hippocampal volume. Points and bars show three-way interaction coefficients and 95% confidence intervals. Negative coefficients indicate a more negative association among NHA than NHW participants. *P* values were adjusted for false discovery rate across the three outcomes. **b**, Race-stratified associations between baseline frontal juxtacortical WMH burden (layer 4) and annual percentage change in AD-signature cortical thickness in EHBS. Linear mixed-effects models were adjusted for age, sex, years of education and intracranial volume. EHBS, Emory Healthy Brain Study; NACC, National Alzheimer’s Coordinating Center; NHA, non-Hispanic African American; NHW, non-Hispanic White; WMH, white matter hyperintensities.

In NACC, the racial difference in the association between baseline frontal juxtacortical WMH burden (layer 4) and subsequent AD-signature cortical thinning was directionally consistent with EHBS but did not reach statistical significance (β = −1.01 × 10^−3^ per year, 95% CI −2.04 × 10^−3^ to 0.02 × 10^−3^, unadjusted *P* = 0.054, FDR-adjusted *P* = 0.163) (Fig. 6a).

## 3. Discussion

In this multi-cohort study of cognitively unimpaired older adults, greater WMH burden was more strongly associated with poorer verbal memory among NHA than NHW participants in EHBS, with independent confirmation in NACC. These racial differences were most evident in frontal juxtacortical white matter (layer 4) in EHBS and parietal deep white matter (layer 3) in NACC and did not simply reflect the broader spatial distribution of WMH burden. The estimates changed little after adjustment for cohort-specific AD biomarkers, vascular risk factors, structural imaging measures and *APOE* genotype. Adjustment for neighborhood deprivation in EHBS produced a similar result. Generalized random forests further supported this pattern without relying on a linear interaction model. In longitudinal analyses, higher frontal juxtacortical WMH burden was associated with faster subsequent AD-signature cortical thinning among NHA than NHW participants in EHBS, while the interaction in NACC was in the same direction.

Previous studies examining racial differences in the association between WMH burden and cognition have yielded inconsistent findings^4–9^. Most have characterized WMH using total or global burden, which could obscure spatial variation in underlying mechanisms and cognitive associations^10,13,14^. By resolving WMH by depth and lobe, we identified race-related effects localized to frontal juxtacortical white matter in EHBS and parietal deep white matter in NACC. Notably, the regional pattern of these effects did not mirror that of racial differences in WMH burden. This spatial dissociation indicates that regional cognitive vulnerability is unlikely to be explained solely by differences in WMH burden. The lobar localization also varied between cohorts, potentially reflecting differences in verbal memory assessment. RAVLT word-list recall places greater demands on executive organization than story recall^16,17^, which may increase sensitivity to frontal white matter involvement in EHBS. This regional variation could also be attributed to differences in participant characteristics and variability across NACC centers. Despite this heterogeneity, the findings were broadly consistent between EHBS and NACC.

The longitudinal analyses extend the cross-sectional findings by associating localized WMH burden with later structural change. Previous studies have linked WMH to cognitive decline in cognitively unimpaired older adults and to subsequent cortical thinning, with greater regional effects reported among African American than White older adults^3,18^. In EHBS, the relationship between baseline frontal juxtacortical WMH burden and subsequent AD-signature cortical thinning was more pronounced among NHA than NHW participants, with a similar directional pattern in NACC. The AD-signature cortex encompasses temporal regions characteristically affected in AD^19^. These findings suggest that race-related differences involving regional WMH burden extend beyond verbal memory to subsequent cortical thinning in AD-vulnerable regions.

Accounting for measured vascular risk factors, including hypertension, diabetes, dyslipidemia and obesity, did not substantially alter the racial differences in EHBS or NACC. In EHBS, the difference remained significant after further adjustment for pulse wave velocity. This pattern is consistent with reports that arterial stiffness and related systemic vascular measures are more strongly associated with periventricular than deep WMH in some studies^20,21^, whereas the effect in EHBS was localized to juxtacortical white matter. Vascular influences on WMH are nevertheless spatially heterogeneous, as hypertension and diabetes have also been associated with deep frontal WMH^10^. Moreover, current vascular diagnoses and a single pulse wave velocity measurement may not fully capture cumulative exposure, severity, treatment or control. Therefore, the vascular factors measured here did not adequately explain the observed racial differences, although vascular mechanisms cannot be excluded.

Educational and social exposures are also plausible contributors to the observed differences. Years of schooling provide an incomplete measure of educational experience and quality, which can vary widely among individuals with similar educational attainment^22,23^. In EHBS, receptive vocabulary, a proxy for premorbid ability, attenuated racial differences across several neuropsychological measures to a greater extent than years of education^24^. Social exposures have been associated with faster WMH accumulation among older African Americans^25^, and structural and social determinants of health have been reported to modify associations between WMH and cognition among African Americans^26^. Adjustment for ADI yielded a similar estimate in EHBS, although differential missingness by race limited interpretation. Broader social and environmental exposures accumulated across the life course, beyond those reflected by years of education and neighborhood deprivation, could influence WMH-related cognitive vulnerability^22,27^.

The absence of a corresponding racial difference in HABS-HD demonstrates that effect modification by race is not uniform across cohorts. In a previous study of HABS-HD, greater global WMH burden was associated with poorer memory in both African American and White participants, without evidence that the association varied by race^28^. The lack of a racial difference in the present HABS-HD analysis would similarly reflect an association between WMH burden and memory shared by both groups. NHW participants in HABS-HD had fewer years of education and higher prevalences of hypertension and diabetes than NHW participants in EHBS and NACC, suggesting that variation in educational experience or cumulative vascular exposure could contribute to the cross-cohort heterogeneity. Education can modify WMH-related cognitive decline, although its role in cognitive reserve may differ across racial and ethnic groups^9^. Differences in recruitment, participant characteristics and memory assessment, together with the close alignment of race and scanner platform in HABS-HD, further limit direct comparisons across cohorts.

Several limitations should be considered. First, the observational design may be subject to residual confounding from unmeasured or incompletely characterized vascular, educational, social and environmental exposures. Second, missing AD biomarker, vascular and genotype data reduced the complete-case sample sizes for some NACC sensitivity analyses. Finally, the smaller longitudinal samples limited precision for detecting racial differences in subsequent structural change.

In conclusion, WMH burden showed a greater association with poorer verbal memory among NHA than NHW participants in both EHBS and NACC. These racial differences localized to frontal juxtacortical white matter (layer 4) in EHBS and parietal deep white matter (layer 3) in NACC, rather than predominantly periventricular regions. The cross-sectional findings remained consistent across covariate-adjusted models and were further supported by generalized random forests. Longitudinally, the association between frontal juxtacortical WMH burden and subsequent AD-signature cortical thinning was stronger among NHA than NHW participants in EHBS, with the interaction in NACC showing the same direction. Spatially resolved WMH measures could therefore provide insight into early heterogeneity in the vascular contributions to cognitive aging.

## 4. Methods

### 4.1 Participants

EHBS is a longitudinal study of community-dwelling adults recruited through the Emory Healthy Aging Study registry^29^. NACC compiles standardized clinical, neuropsychological and imaging data from US Alzheimer’s Disease Research Centers^30,31^. The Emory center was excluded from NACC to maintain independence from EHBS. HABS-HD is a longitudinal study of older adults in North Texas that uses community-based participatory recruitment methods^32^ (Fig. 1a).

Analyses were restricted to participants who were cognitively unimpaired at baseline, self-identified as NHA or NHW and had eligible T1-weighted and fluid-attenuated inversion recovery (FLAIR) magnetic resonance imaging (MRI) scans. The earliest eligible MRI scan was designated as baseline, and subsequent eligible MRI scans contributed to longitudinal analyses regardless of later cognitive status. Cohort derivation and analytic samples are shown in Supplementary Fig. 1.

The present analyses were approved by the Emory University Institutional Review Board. All EHBS participants provided written informed consent, and study procedures were conducted in accordance with the Declaration of Helsinki and applicable Health Insurance Portability and Accountability Act requirements. NACC data were collected at participating Alzheimer’s Disease Research Centers under local institutional review board approval, with written informed consent obtained from participants. The HABS-HD protocol was approved by the North Texas Regional Institutional Review Board, and participants provided written informed consent.

### 4.2 MRI acquisition

In EHBS, MRI data were acquired on a Siemens Magnetom Prisma 3T scanner equipped with a 32-channel head array coil. T1-weighted anatomical images were acquired using a magnetization-prepared rapid acquisition with gradient echo (MPRAGE) sequence (TR/TE = 2,300/2.96 ms, TI = 900 ms, FA = 9°, voxel size = 1.0 × 1.0 × 1.0 mm³, 208 slices). FLAIR images were acquired with TR/TE = 4,800/441 ms, TI = 1,650 ms, FA = 120°, voxel size = 1.0 × 1.0 × 1.2 mm³ and 160 slices.

In NACC, MRI data were acquired across 34 centers on 3T Siemens, GE Healthcare and Philips scanners using vendor- and platform-specific SCAN MRI protocols (https://scan.naccdata.org). T1-weighted images predominantly had voxel sizes of 1.0 × 1.0 × 1.0 mm³, 1.05 × 1.05 × 1.0 mm³ or 1.05 × 1.05 × 1.2 mm³. FLAIR images predominantly had voxel sizes of 1.0 × 1.0 × 1.2 mm³ or 1.0 × 1.0 × 1.0 mm³. Sequence timing and other acquisition parameters varied by scanner platform according to the SCAN protocols.

In HABS-HD, MRI data were acquired on Siemens Magnetom Skyra and Vida 3T scanners. T1-weighted anatomical images were acquired using an MPRAGE sequence (TR = 2,150–2,300 ms, TE = 2.91–3.20 ms, TI = 900 ms, FA = 9°), predominantly with voxel sizes of 1.0 × 1.0 × 1.0 mm³ or 1.05 × 1.05 × 1.2 mm³. FLAIR images were acquired using two protocols: TR/TE = 4,800/441 ms, TI = 1,650 ms, FA = 120° and voxel size = 1.0 × 1.0 × 1.2 mm³; or TR/TE = 5,000/393 ms, TI = 1,700 ms, FA = 120° and voxel size = 1.0 × 1.0 × 1.0 mm³.

### 4.3 MRI processing

The same image processing workflow was applied across cohorts. T1-weighted and FLAIR images were separately corrected for intensity non-uniformity using N4 bias field correction in ANTs 2.2 (https://github.com/ANTsX/ANTs). Bias-corrected T1-weighted images were then rigidly registered to the corresponding FLAIR images. WMH were segmented from the bias-corrected FLAIR and co-registered T1-weighted images using the Emory Robust WMH Segmentation pipeline^33^, implemented with nnU-Net^34^.

T1-weighted images were processed with FreeSurfer 7.4^35^ to reconstruct cortical surfaces and derive Desikan–Killiany regional cortical thickness and surface area, hippocampal volume and estimated intracranial volume. AD-signature cortical thickness was calculated as the bilateral, surface area-weighted mean thickness of the entorhinal, fusiform, inferior temporal and middle temporal regions^19^. Matched-lobe cortical thickness was calculated using the same surface-area weighting across regions within the corresponding lobe. Hippocampal volume, intracranial volume, AD-signature cortical thickness and matched-lobe cortical thickness were log_10_ transformed before analysis.

Cerebral white matter was divided into frontal, parietal, temporal and occipital sectors crossed with four participant-normalized depth layers using the bullseye parcellation^36^. The depth layers were defined relative to the ventricular and cortical surfaces, with layer 1 representing periventricular white matter, layers 2–3 deep white matter and layer 4 juxtacortical white matter. Bilateral WMH volumes were calculated for the 16 lobe-by-layer compartments in native FLAIR space. WMH volumes were log_10_ transformed after adding 1 mm³ to accommodate zero values.

All registrations, cortical reconstructions, parcellations and WMH segmentations underwent visual quality control. Scans with major imaging abnormalities, artefacts or processing failures were excluded (Supplementary Fig. 1).

### 4.4 Neuropsychological assessments

EHBS participants completed a neuropsychological test battery that included the Rey Auditory Verbal Learning Test (RAVLT), Rey Complex Figure Test, Number Span Test, Judgment of Line Orientation, Trail Making Test Parts A and B, Letter Fluency and Animal Fluency. Cognitive domain scores for verbal memory, visual memory, working memory, executive function and processing speed, and language and fluency were derived using exploratory factor analysis^37^.

For NACC, neuropsychological measures included individual tests from the Uniform Data Set battery and harmonized, co-calibrated domain scores. Craft Story 21 delayed recall, Benson Complex Figure delayed recall and Number Span Backward were used to assess verbal memory, visual memory and working memory, respectively^38^. Harmonized executive function and language scores from the Alzheimer’s Disease Sequencing Project Phenotype Harmonization Consortium were used for the corresponding domains^39^.

For HABS-HD, Spanish-English Verbal Learning Test (SEVLT) delayed recall, Wechsler Memory Scale–Third Edition Digit Span Total, Digit Symbol Substitution and Animal Naming were used to assess verbal memory, working memory, executive function and processing speed, and language and fluency, respectively^32^.

The cohort-specific delayed verbal recall measures were RAVLT delayed recall in EHBS, Craft Story delayed recall in NACC and SEVLT delayed recall in HABS-HD. RAVLT and SEVLT are word-list measures, whereas Craft Story is a narrative recall measure. All neuropsychological scores included in the analyses were standardized as z scores within cohort.

### 4.5 Alzheimer’s disease biomarkers

In EHBS, CSF was collected using standardized pre-analytical procedures, aliquoted within 1 h of collection and stored at −80 °C. CSF amyloid-β 1–42 (Aβ42), total tau and tau phosphorylated at threonine 181 (p-tau181) were measured in a single laboratory in the Emory Goizueta Alzheimer’s Clinical Research Unit using Elecsys immunoassays on a Roche Cobas e601 analyzer. The p-tau181/Aβ42 ratio was calculated as a combined measure of amyloid and phosphorylated tau burden and was log_10_ transformed before analysis.

In NACC and HABS-HD, amyloid PET burden was quantified on the Centiloid scale^40^. Centiloid measures in NACC were obtained from the Alzheimer’s Disease Sequencing Project Phenotype Harmonization Consortium release; in HABS-HD, amyloid PET was acquired with [^18^F]florbetaben^32^.

### 4.6 Statistical analysis

Cross-sectional associations between WMH burden and neuropsychological performance were evaluated using linear regression models including race, WMH burden and their interaction, with age, sex, years of education and intracranial volume as covariates. Models were fitted by ordinary least squares with heteroskedasticity-consistent (HC3) standard errors, with NHW participants as the reference group. Scanner manufacturer was additionally included as a fixed effect in NACC. Scanner platform was not included in HABS-HD because of near collinearity with race. Spatial localization followed a prespecified two-stage hierarchy in which the four white matter depth layers were tested first, followed by the four lobar sectors within significant depth layers.

Robustness was assessed by additionally adjusting the localized associations for a cohort-specific AD biomarker, vascular risk factors (hypertension, diabetes, dyslipidemia and obesity), both the AD biomarker and vascular risk factors, AD-signature cortical thickness, hippocampal volume or *APOE* ε4. In EHBS, sensitivity analyses additionally adjusted for Area Deprivation Index (ADI) or for pulse wave velocity together with vascular risk factors. ADI was not evaluated in NACC because data were available for only 33% of participants, and availability varied substantially by race and center. For each covariate-adjusted analysis, the primary model was refitted in the same complete-case sample. Sensitivity to acquisition and center effects in NACC was assessed using seven specifications: no acquisition adjustment, scanner manufacturer as a fixed effect, scanner manufacturer and FLAIR slice thickness as fixed effects, scanner manufacturer with standard errors clustered by center, center fixed effects, a center random intercept, or a center–scanner-model random intercept. Leave-one-center-out analyses were also performed to assess sensitivity to individual centers.

Generalized random forests with 4,000 trees^41^ were used to evaluate the association between WMH burden and delayed recall without imposing a linear interaction between race and WMH burden. The same WMH exposure and core covariates were used as in the linear regression models, including scanner manufacturer in NACC. Participant-level conditional effects and doubly robust average partial effects within racial groups were estimated, and the between-group difference was estimated by best linear projection of the participant-level effects onto race.

Longitudinal associations between baseline frontal juxtacortical WMH burden (layer 4) and subsequent structural change were evaluated using linear mixed-effects models with participant-specific random intercepts and slopes for time. Models included the interaction among race, baseline WMH burden and time and were fitted separately for matched frontal cortical thickness, AD-signature cortical thickness and hippocampal volume, with baseline age, sex, years of education and intracranial volume as covariates. In NACC, scanner manufacturer did not vary within participants across visits and was therefore not included as a covariate.

For interaction analyses, *P* values were adjusted using the Benjamini–Hochberg false discovery rate (FDR) procedure^42^ separately within each prespecified testing family: the four depth layers for each neuropsychological measure, the four lobes within each depth-specific analysis, including each covariate-adjusted model, and the three longitudinal structural measures. Race-stratified follow-up analyses were not included in these multiple-testing corrections. All tests were two-sided, with *P* < 0.05 considered statistically significant. Analyses were conducted in R 4.6.

## Acknowledgements

The NACC database is funded by NIA/NIH Grant U24 AG072122. NACC data are contributed by the NIA-funded ADRCs: P30 AG062429 (PI James Brewer, MD, PhD), P30 AG066468 (PI Oscar Lopez, MD), P30 AG062421 (PI Teresa Gomez-Isla, MD), P30 AG066509 (PI Thomas Grabowski, MD), P30 AG066514 (PI Mary Sano, PhD), P30 AG066530 (PIs Helena Chui, MD; Arthur Toga, PhD), P30 AG066507 (PI Marilyn Albert, PhD), P30 AG066444 (PI David Holtzman, MD), P30 AG066518 (PIs Lisa Silbert, MD; Kevin Duff, PhD), P30 AG066512 (PI Thomas Wisniewski, MD), P30 AG066462 (PI Scott Small, MD), P30 AG072979 (PI David Wolk, MD), P30 AG072972 (PIs Charles DeCarli, MD; Rachel Whitmer, PhD), P30 AG072976 (PI Andrew Saykin, PsyD), P30 AG072975 (PI Julie Schneider, MD, MS), P30 AG072978 (PI Ann McKee, MD), P30 AG072977 (PI Robert Vassar, PhD), P30 AG066519 (PI Joshua Grill, PhD), P30 AG062677 (PIs Brad Boeve, MD; Ronald Petersen, MD, PhD), P30 AG079280 (PI Jessica Langbaum, PhD), P30 AG062422 (PI Gil Rabinovici, MD), P30 AG066511 (PI Allan Levey, MD, PhD), P30 AG072946 (PI Linda Van Eldik, PhD), P30 AG062715 (PI Sanjay Asthana, MD, FRCP), P30 AG072973 (PI Russell Swerdlow, MD), P30 AG066506 (PIs Glenn Smith, PhD, ABPP; David Lowenstein, PhD; Ranjan Duara, MD), P30 AG066508 (PIs Stephen Strittmatter, MD, PhD; Christopher Van Dyck, MD), P30 AG066515 (PI Victor Henderson, MD, MS), P30 AG072947 (PI Suzanne Craft, PhD), P30 AG072931 (PI Henry Paulson, MD, PhD), P30 AG066546 (PIs Sudha Seshadri, MD; Gladys Maestre, MD, PhD), P30 AG086401 (PI Erik Roberson, MD, PhD), P30 AG086404 (PI Gary Rosenberg, MD), P30 AG086403 (PI Angela Jefferson, PhD), P30 AG072958 (PIs Heather Whitson, MD; Gwenn Garden, MD, PhD), P30 AG072959 (PI Jagan Pillai, MD, PhD), P30 AG092752 (PI Ihab Hajjar, MD, MS).

SCAN is a multi-institutional project funded by the National Institute on Aging through grant U24 AG067418, awarded in May 2020. Data collected by SCAN and shared by NACC are contributed by the NIA-funded ADRCs as follows: Arizona Alzheimer’s Center – P30 AG072980 (PI Eric Reiman, MD); R01 AG069453 (contact PI Eric Reiman, MD); P30 AG019610 (PI Eric Reiman, MD); and the State of Arizona, which provided additional funding supporting the center; Boston University – P30 AG013846 (PI Neil Kowall, MD); Cleveland ADRC – P30 AG062428 (PI James Leverenz, MD); Cleveland Clinic, Las Vegas – P20 AG068053; Columbia – P50 AG008702 (PI Scott Small, MD); Duke/UNC ADRC – P30 AG072958; Emory University – P30 AG066511 (PI Allan Levey, MD, PhD); Indiana University – R01 AG19771 (PI Andrew Saykin, PsyD); P30 AG10133 (PI Andrew Saykin, PsyD); P30 AG072976 (PI Andrew Saykin, PsyD); R01 AG061788 (PI Shannon Risacher, PhD); R01 AG053993 (PI Yu-Chien Wu, MD, PhD); U01 AG057195 (PI Liana Apostolova, MD); U19 AG063911 (PI Bradley Boeve, MD); and the Indiana University Department of Radiology and Imaging Sciences; Johns Hopkins – P30 AG066507 (PI Marilyn Albert, PhD); Mayo Clinic – P50 AG016574 (PI Ronald Petersen, MD, PhD); Mount Sinai – P30 AG066514 (PI Mary Sano, PhD); R01 AG054110 (PI Trey Hedden, PhD); R01 AG053509 (PI Trey Hedden, PhD); New York University – P30 AG066512-01S2 (PI Thomas Wisniewski, MD); R01 AG056031 (PI Ricardo Osorio, MD); R01 AG056531 (PIs Ricardo Osorio, MD; Girardin Jean-Louis, PhD); Northwestern University – P30 AG013854 (PI Robert Vassar, PhD); R01 AG045571 (PI Emily Rogalski, PhD); R56 AG045571 (PI Emily Rogalski, PhD); R01 AG067781 (PI Emily Rogalski, PhD); U19 AG073153 (PI Emily Rogalski, PhD); R01 DC008552 (PI M.-Marsel Mesulam, MD); R01 AG077444 (PIs M.-Marsel Mesulam, MD; Emily Rogalski, PhD); R01 NS075075 (PI Emily Rogalski, PhD); R01 AG056258 (PI Emily Rogalski, PhD); Oregon Health and Science University – P30 AG066518 (PIs Lisa Silbert, MD; Kevin Duff, PhD); R56 AG074321 (PI Jeffrey Kaye, MD); Rush University – P30 AG010161 (PI David Bennett, MD); Stanford – P30 AG066515; P50 AG047366 (PI Victor Henderson, MD, MS); University of Alabama, Birmingham – P20; University of California, Davis – P30 AG10129 (PI Charles DeCarli, MD); P30 AG072972 (PI Charles DeCarli, MD); University of California, Irvine – P50 AG016573 (PI Frank LaFerla, PhD); University of California, San Diego – P30 AG062429 (PI James Brewer, MD, PhD); University of California, San Francisco – P30 AG062422 (PI Gil D. Rabinovici, MD); University of Kansas – P30 AG035982 (PI Russell Swerdlow, MD); University of Kentucky – P30 AG028283-15S1 (PIs Linda Van Eldik, PhD; Brian Gold, PhD); University of Michigan ADRC – P30 AG053760 (PI Henry Paulson, MD, PhD); P30 AG072931 (PI Henry Paulson, MD, PhD); Cure Alzheimer’s Fund 200775 (PI Henry Paulson, MD, PhD); U19 NS120384 (PI Charles DeCarli, MD; University of Michigan site PI Henry Paulson, MD, PhD); R01 AG068338 (MPIs Bruno Giordani, PhD; Carol Persad, PhD; Yi Murphey, PhD); S10 OD026738-01 (PI Douglas Noll, PhD); R01 AG058724 (PI Benjamin Hampstead, PhD); R35 AG072262 (PI Benjamin Hampstead, PhD); W81XWH2110743 (PI Benjamin Hampstead, PhD); R01 AG073235 (PI Nancy Chiaravalloti; University of Michigan site PI Benjamin Hampstead, PhD); 1I01RX001534 (PI Benjamin Hampstead, PhD); IRX001381 (PI Benjamin Hampstead, PhD); University of New Mexico – P20 AG068077 (PI Gary Rosenberg, MD); University of Pennsylvania – State of PA project 2019NF4100087335 (PI David Wolk, MD); Rooney Family Research Fund (PI David Wolk, MD); R01 AG055005 (PI David Wolk, MD); University of Pittsburgh – P50 AG005133 (PI Oscar Lopez, MD); University of Southern California – P50 AG005142 (PI Helena Chui, MD); University of Washington – P50 AG005136 (PI Thomas Grabowski, MD); University of Wisconsin – P50 AG033514 (PI Sanjay Asthana, MD, FRCP); Vanderbilt University – P20 AG068082; Wake Forest – P30 AG072947 (PI Suzanne Craft, PhD); Washington University, St. Louis – P01 AG03991 (PI John Morris, MD); P01 AG026276 (PI John Morris, MD); P20 MH071616 (PI Dan Marcus); P30 AG066444 (PI John Morris, MD); P30 NS098577 (PI Dan Marcus); R01 AG021910 (PI Randy Buckner); R01 AG043434 (PI Catherine Roe); R01 EB009352 (PI Dan Marcus); UL1 TR000448 (PI Brad Evanoff); U24 RR021382 (PI Bruce Rosen); Avid Radiopharmaceuticals/Eli Lilly; Yale – P50 AG047270 (PI Stephen Strittmatter, MD, PhD); R01 AG052560 (MPIs Christopher van Dyck, MD; Richard Carson, PhD); R01 AG062276 (PI Christopher van Dyck, MD); 1Florida – P30 AG066506-03 (PI Glenn Smith, PhD); P50 AG047266 (PI Todd Golde, MD, PhD).

Data used in this study were obtained in part from HABS-HD. Research reported in this publication was supported by the National Institute on Aging of the National Institutes of Health under Award Numbers R01AG054073, R01AG058533, R01AG070862, P41EB015922 and U19AG078109. The content is solely the responsibility of the authors and does not necessarily represent the official views of the National Institutes of Health.

## Data availability

EHBS data supporting the findings of this study are available to qualified investigators upon reasonable request, subject to applicable ethical requirements and a data use agreement with the authors’ institution. Data from NACC (https://www.naccdata.org) and HABS-HD (https://apps.unthsc.edu/itr/data) are available upon request, subject to their respective data use agreements.

## Author contributions

J.W., A.I.L., J.J.L., and D.Q. conceptualized the study and interpreted the data. J.W., Q.L., and T.A.J. performed the acquisition of the data. J.W., Q.L., and T.A.J. performed the analysis and quality control of the image data. J.W., J.J.L., and D.Q. wrote the manuscript and all authors contributed to the reviewing and editing the manuscript.

## Supplementary materials

**Supplementary Fig. 1.**
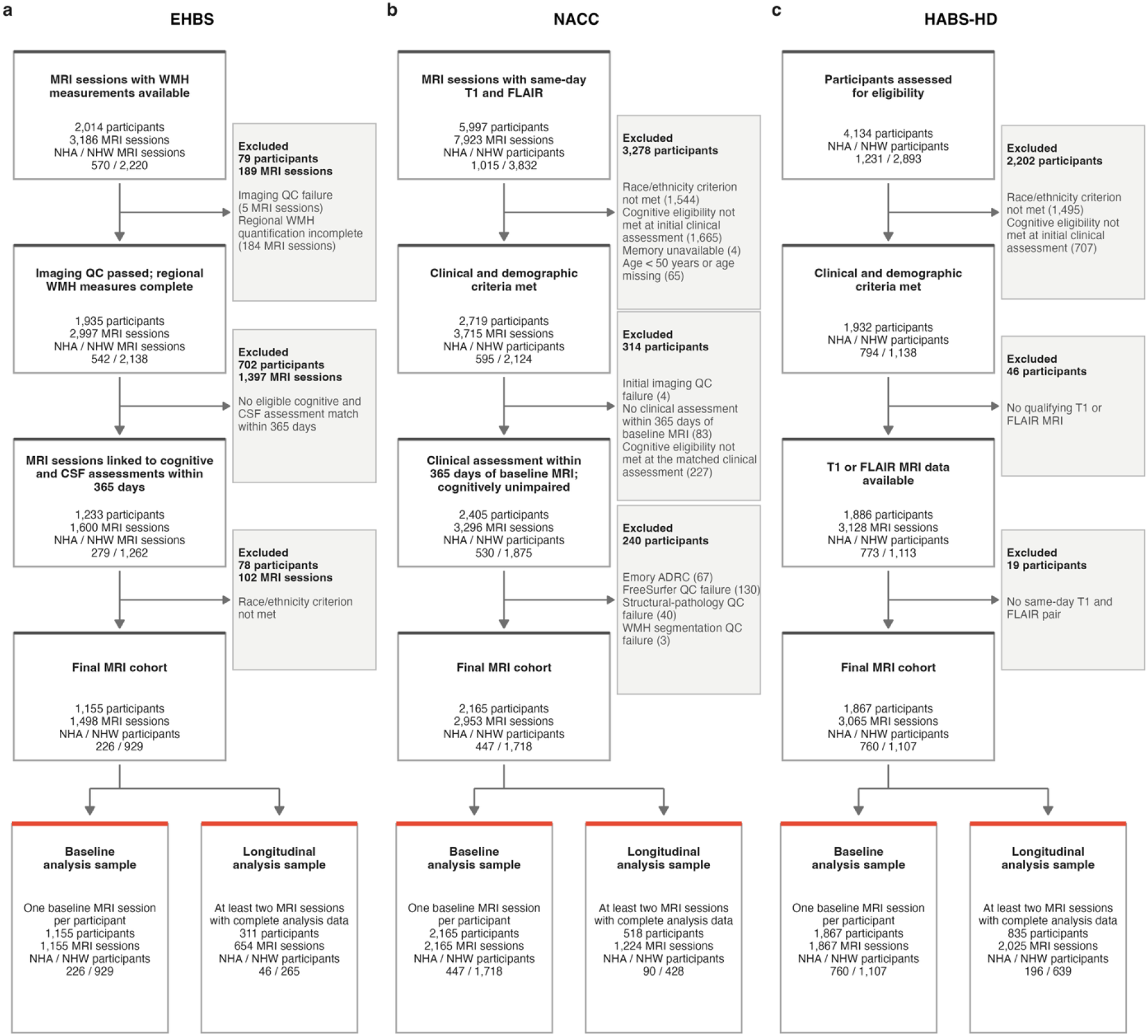
Cohort derivation and analytic samples. **a**, EHBS. **b**, NACC. **c**, HABS-HD. Flowcharts show participant inclusion and exclusion for each cohort, with retained samples in white and exclusions in grey. The earliest eligible MRI was defined as baseline, and subsequent eligible scans contributed to longitudinal analyses. Final analytic samples included NHA and NHW participants. Emory participants were excluded from NACC to maintain independence from EHBS. ADRC, Alzheimer’s Disease Research Center; CSF, cerebrospinal fluid; EHBS, Emory Healthy Brain Study; FLAIR, fluid-attenuated inversion recovery; HABS-HD, Health and Aging Brain Study–Health Disparities; MRI, magnetic resonance imaging; NACC, National Alzheimer’s Coordinating Center; NHA, non-Hispanic African American; NHW, non-Hispanic White; QC, quality control; WMH, white matter hyperintensities.

**Supplementary Fig. 2.**
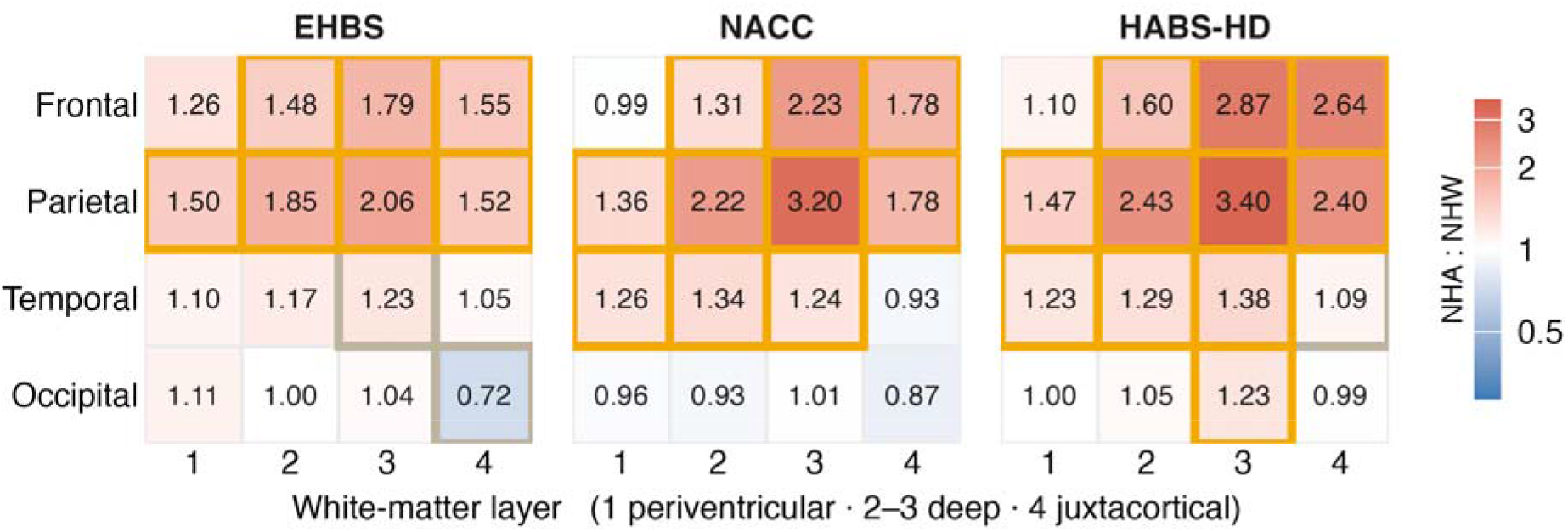
Racial differences in WMH burden across lobes and white matter depth. Values are 10^β^, the back-transformed race coefficients from linear regression models of log_10_(WMH volume + 1 mm³), and represent adjusted NHA-to-NHW ratios on the shifted WMH scale. Values greater than 1 indicate higher WMH burden among NHA participants. Orange and grey outlines indicate *P* values < 0.05 and 0.05–0.10, respectively, after false discovery rate correction. Linear regression models were adjusted for age, sex, years of education and intracranial volume, with scanner manufacturer additionally included as a fixed effect in NACC. Heteroskedasticity-consistent (HC3) standard errors were used. EHBS, Emory Healthy Brain Study; HABS-HD, Health and Aging Brain Study–Health Disparities; NACC, National Alzheimer’s Coordinating Center; NHA, non-Hispanic African American; NHW, non-Hispanic White; WMH, white matter hyperintensities.

**Supplementary Fig. 3.**
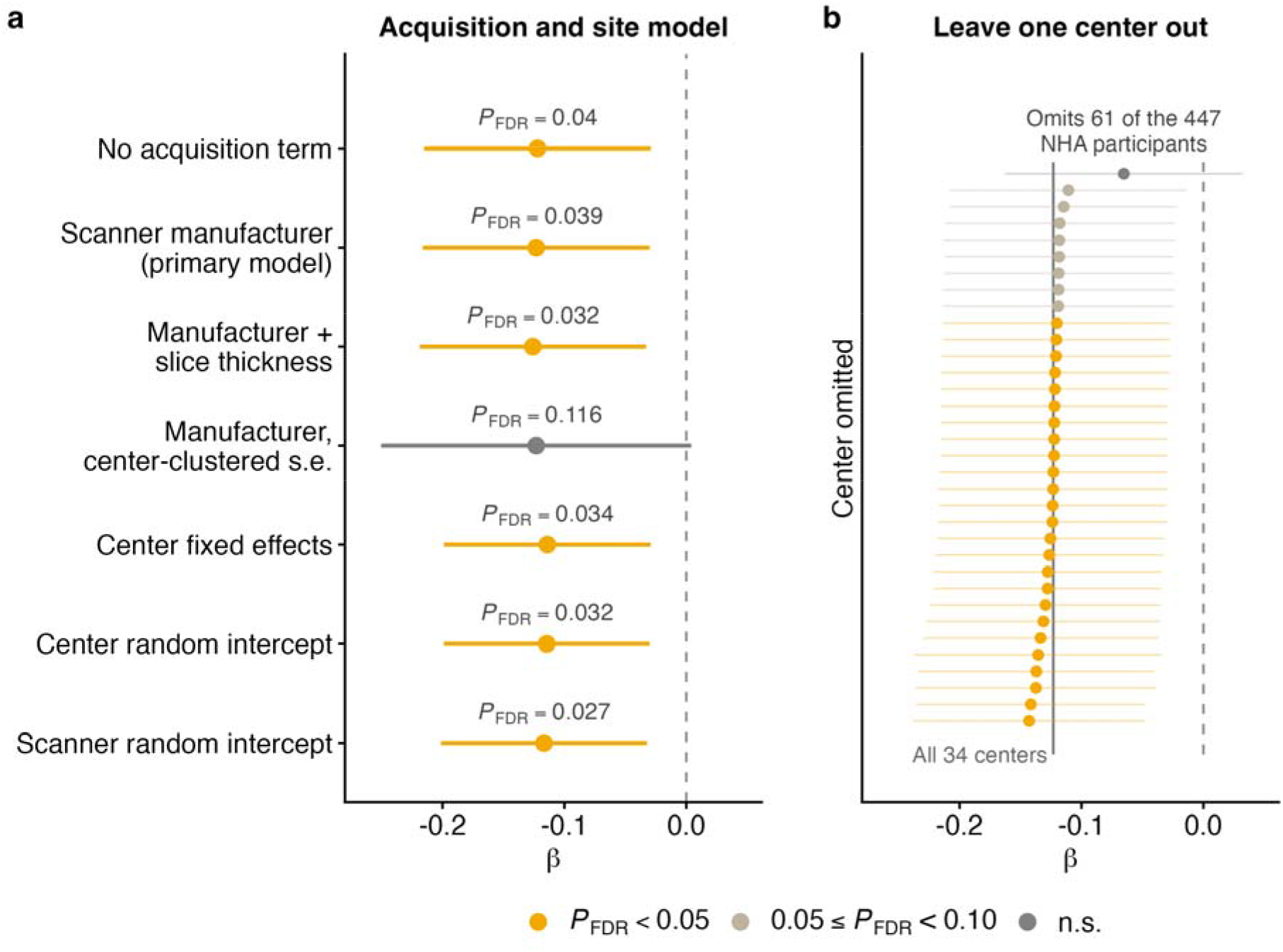
Sensitivity of racial differences to acquisition and center specifications in NACC. **a**, Estimates from seven acquisition and center specifications: no acquisition adjustment, scanner manufacturer (primary model), scanner manufacturer plus slice thickness, scanner manufacturer with center-clustered standard errors, center fixed effects, center random intercept and center–scanner-model random intercept. **b**, Estimates from 34 analyses in which each center was excluded in turn; the vertical line indicates the full-sample estimate. Both panels show the interaction between race and parietal layer 3 WMH burden for Craft Story delayed recall. Points and bars show interaction coefficients and 95% confidence intervals. Models were adjusted for age, sex, years of education and intracranial volume. *P* values were adjusted for false discovery rate. NACC, National Alzheimer’s Coordinating Center; NHA, non-Hispanic African American; WMH, white matter hyperintensities.

**Supplementary Table 1.**
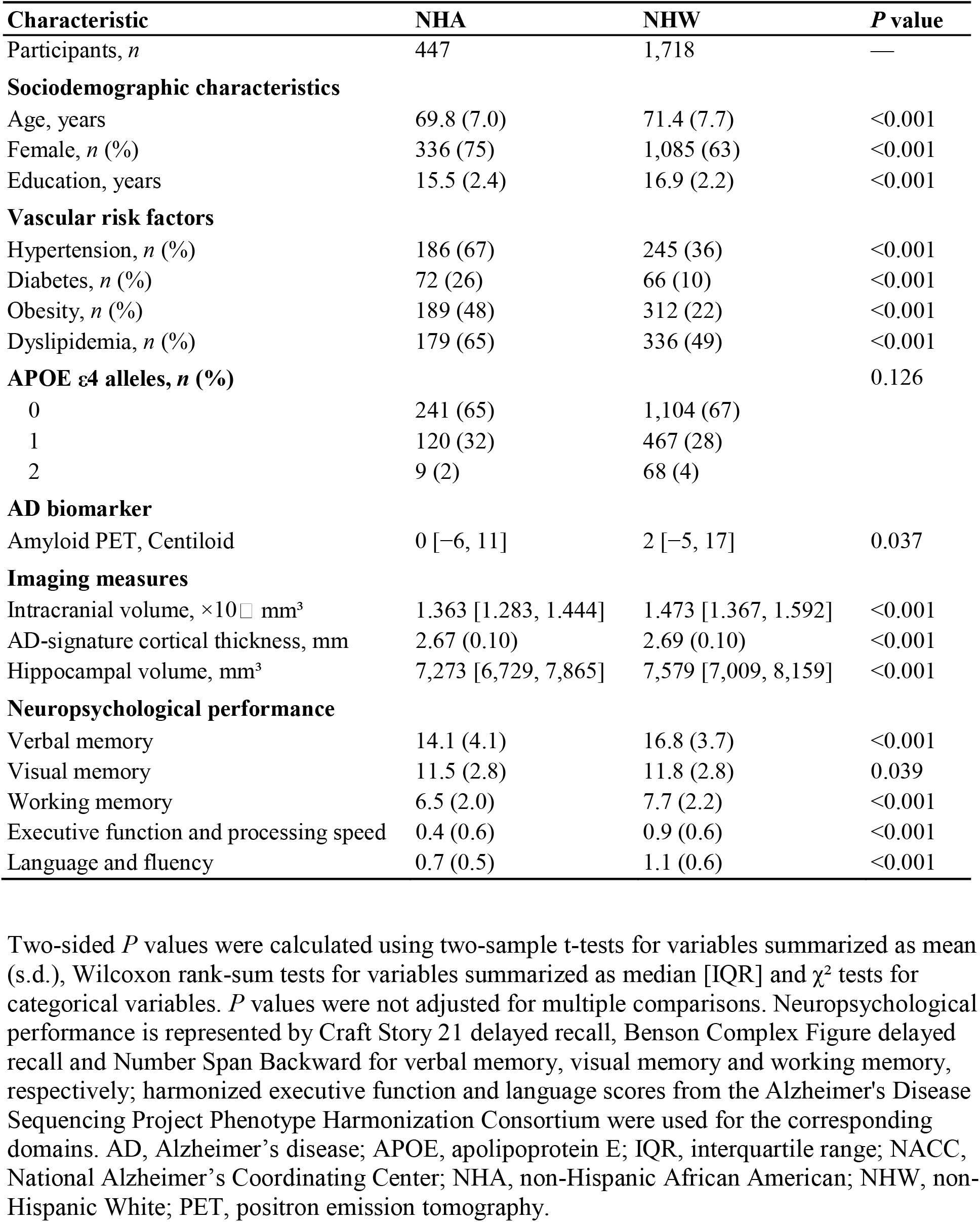
Participant characteristics in NACC.

| Characteristic | NHA | NHW | *P* value |
|---|---|---|---|
| Participants, *n* | 447 | 1,718 | — |
| **Sociodemographic characteristics** |  |  |  |
| Age, years | 69.8 (7.0) | 71.4 (7.7) | <0.001 |
| Female, *n* (%) | 336 (75) | 1,085 (63) | <0.001 |
| Education, years | 15.5 (2.4) | 16.9 (2.2) | <0.001 |
| **Vascular risk factors** |  |  |  |
| Hypertension, *n* (%) | 186 (67) | 245 (36) | <0.001 |
| Diabetes, *n* (%) | 72 (26) | 66 (10) | <0.001 |
| Obesity, *n* (%) | 189 (48) | 312 (22) | <0.001 |
| Dyslipidemia, *n* (%) | 179 (65) | 336 (49) | <0.001 |
| **APOE ε4 alleles, *n* (%)** |  |  | 0.126 |
| 0 | 241 (65) | 1,104 (67) |  |
| 1 | 120 (32) | 467 (28) |  |
| 2 | 9 (2) | 68 (4) |  |
| **AD biomarker** |  |  |  |
| Amyloid PET, Centiloid | 0 [−6, 11] | 2 [−5, 17] | 0.037 |
| **Imaging measures** |  |  |  |
| Intracranial volume, ×10□ mm³ | 1.363 [1.283, 1.444] | 1.473 [1.367, 1.592] | <0.001 |
| AD-signature cortical thickness, mm | 2.67 (0.10) | 2.69 (0.10) | <0.001 |
| Hippocampal volume, mm³ | 7,273 [6,729, 7,865] | 7,579 [7,009, 8,159] | <0.001 |
| **Neuropsychological performance** |  |  |  |
| Verbal memory | 14.1 (4.1) | 16.8 (3.7) | <0.001 |
| Visual memory | 11.5 (2.8) | 11.8 (2.8) | 0.039 |
| Working memory | 6.5 (2.0) | 7.7 (2.2) | <0.001 |
| Executive function and processing speed | 0.4 (0.6) | 0.9 (0.6) | <0.001 |
| Language and fluency | 0.7 (0.5) | 1.1 (0.6) | <0.001 |

Two-sided *P* values were calculated using two-sample t-tests for variables summarized as mean (s.d.), Wilcoxon rank-sum tests for variables summarized as median [IQR] and χ² tests for categorical variables. *P* values were not adjusted for multiple comparisons. Neuropsychological performance is represented by Craft Story 21 delayed recall, Benson Complex Figure delayed recall and Number Span Backward for verbal memory, visual memory and working memory, respectively; harmonized executive function and language scores from the Alzheimer's Disease Sequencing Project Phenotype Harmonization Consortium were used for the corresponding domains. AD, Alzheimer’s disease; APOE, apolipoprotein E; IQR, interquartile range; NACC, National Alzheimer’s Coordinating Center; NHA, non-Hispanic African American; NHW, non-Hispanic White; PET, positron emission tomography.

**Supplementary Table 2.**
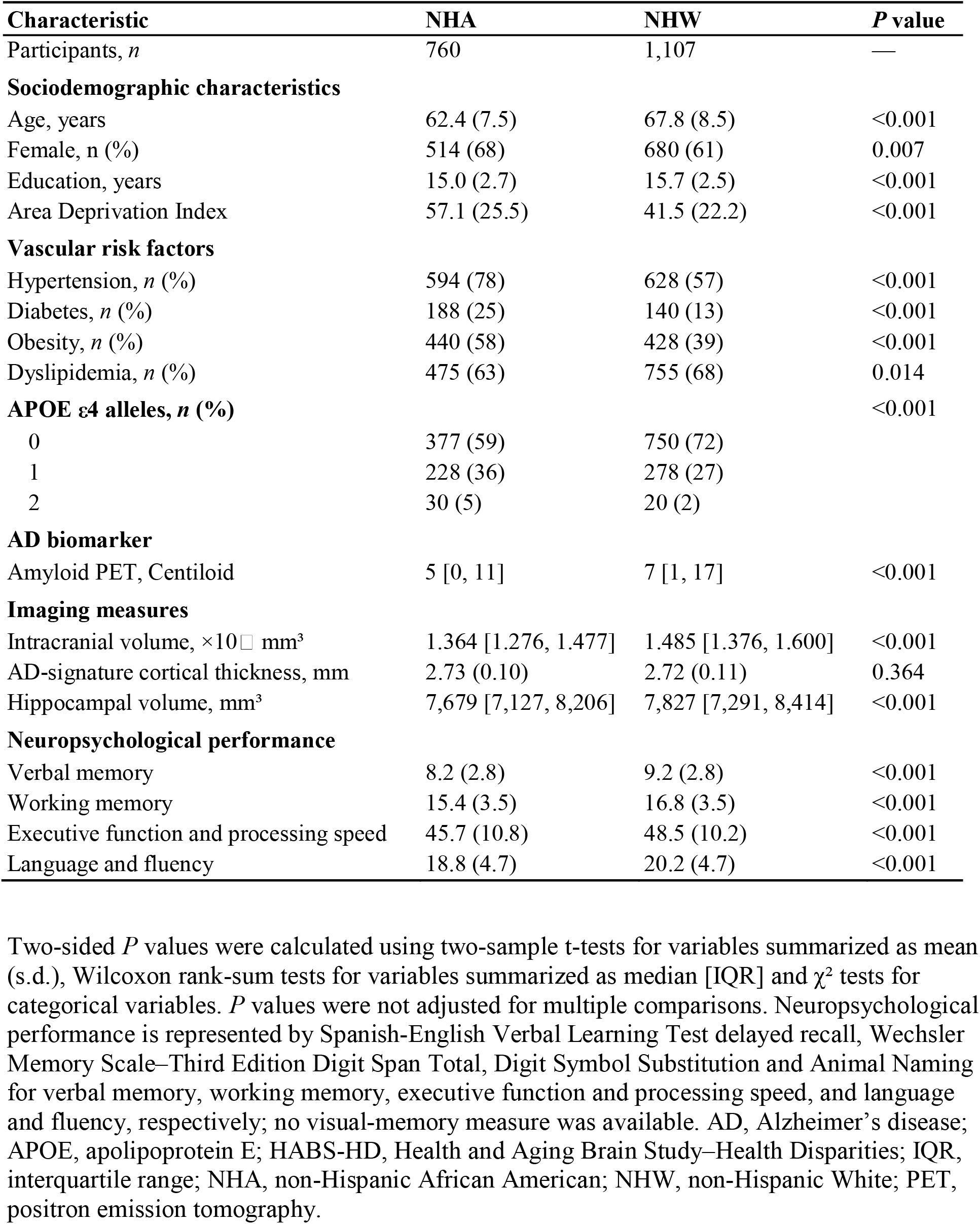
Participant characteristics in HABS-HD.

| Characteristic | NHA | NHW | *P* value |
|---|---|---|---|
| Participants, *n* | 760 | 1,107 | — |
| **Sociodemographic characteristics** |  |  |  |
| Age, years | 62.4 (7.5) | 67.8 (8.5) | <0.001 |
| Female, n (%) | 514 (68) | 680 (61) | 0.007 |
| Education, years | 15.0 (2.7) | 15.7 (2.5) | <0.001 |
| Area Deprivation Index | 57.1 (25.5) | 41.5 (22.2) | <0.001 |
| **Vascular risk factors** |  |  |  |
| Hypertension, *n* (%) | 594 (78) | 628 (57) | <0.001 |
| Diabetes, *n* (%) | 188 (25) | 140 (13) | <0.001 |
| Obesity, *n* (%) | 440 (58) | 428 (39) | <0.001 |
| Dyslipidemia, *n* (%) | 475 (63) | 755 (68) | 0.014 |
| **APOE ε4 alleles, *n* (%)** |  |  | <0.001 |
| 0 | 377 (59) | 750 (72) |  |
| 1 | 228 (36) | 278 (27) |  |
| 2 | 30 (5) | 20 (2) |  |
| **AD biomarker** |  |  |  |
| Amyloid PET, Centiloid | 5 [0, 11] | 7 [1, 17] | <0.001 |
| **Imaging measures** |  |  |  |
| Intracranial volume, ×10□ mm<sup>3</sup> | 1.364 [1.276, 1.477] | 1.485 [1.376, 1.600] | <0.001 |
| AD-signature cortical thickness, mm | 2.73 (0.10) | 2.72 (0.11) | 0.364 |
| Hippocampal volume, mm<sup>3</sup> | 7,679 [7,127, 8,206] | 7,827 [7,291, 8,414] | <0.001 |
| **Neuropsychological performance** |  |  |  |
| Verbal memory | 8.2 (2.8) | 9.2 (2.8) | <0.001 |
| Working memory | 15.4 (3.5) | 16.8 (3.5) | <0.001 |
| Executive function and processing speed | 45.7 (10.8) | 48.5 (10.2) | <0.001 |
| Language and fluency | 18.8 (4.7) | 20.2 (4.7) | <0.001 |

Two-sided *P* values were calculated using two-sample t-tests for variables summarized as mean (s.d.), Wilcoxon rank-sum tests for variables summarized as median [IQR] and χ<sup>2</sup> tests for categorical variables. *P* values were not adjusted for multiple comparisons. Neuropsychological performance is represented by Spanish-English Verbal Learning Test delayed recall, Wechsler Memory Scale–Third Edition Digit Span Total, Digit Symbol Substitution and Animal Naming for verbal memory, working memory, executive function and processing speed, and language and fluency, respectively; no visual-memory measure was available. AD, Alzheimer's disease; APOE, apolipoprotein E; HABS-HD, Health and Aging Brain Study–Health Disparities; IQR, interquartile range; NHA, non-Hispanic African American; NHW, non-Hispanic White; PET, positron emission tomography.

**Supplementary Table 3.** Sensitivity of racial differences to additional covariate adjustment.

| Model | *n* (NHA) | Primary model β (95% CI) | Adjusted model β (95% CI) | Change in β (%) | *P* value |
|---|---|---|---|---|---|
| **EHBS** |  |  |  |  |  |
| Primary | 1,092 (200) | −0.189 (−0.320 to −0.058) | −0.189 (−0.320 to −0.058) | — | 0.019 |
| Primary + Area Deprivation Index | 923 (154) | −0.152 (−0.303 to −0.002) | −0.152 (−0.302 to −0.001) | −0.3 | 0.146 |
| Primary + vascular risk factors & arterial stiffness | 997 (174) | −0.184 (−0.320 to −0.048) | −0.183 (−0.320 to −0.047) | −0.6 | 0.035 |
| **NACC** |  |  |  |  |  |
| Primary | 2,126 (443) | −0.123 (−0.216 to −0.030) | −0.123 (−0.216 to −0.030) | — | 0.039 |
| Primary + AD biomarker | 727 (134) | −0.135 (−0.305 to 0.035) | −0.143 (−0.313 to 0.027) | 5.8 | 0.273 |
| Primary + vascular risk factors | 863 (247) | −0.193 (−0.333 to −0.053) | −0.195 (−0.335 to −0.054) | 1.1 | 0.027 |
| Primary + AD biomarker & vascular risk factors | 249 (66) | −0.218 (−0.555 to 0.120) | −0.205 (−0.533 to 0.124) | −5.9 | 0.736 |
| Primary + AD-signature cortical thickness | 2,126 (443) | −0.123 (−0.216 to −0.030) | −0.123 (−0.217 to −0.030) | 0.2 | 0.038 |
| Primary + hippocampus volume | 2,126 (443) | −0.123 (−0.216 to −0.030) | −0.125 (−0.218 to −0.032) | 1.3 | 0.035 |
| Primary + *APOE* ε4 | 1,970 (366) | −0.100 (−0.201 to 0.001) | −0.102 (−0.202 to −0.001) | 1.3 | 0.194 |

Models examined racial differences in the association between frontal juxtacortical WMH burden (layer 4) and Rey Auditory Verbal Learning Test delayed recall in EHBS and between parietal deep WMH burden (layer 3) and Craft Story 21 delayed recall in NACC. The primary linear regression models were adjusted for age, sex, years of education and intracranial volume, with scanner manufacturer additionally included as a fixed effect in NACC. For each adjusted analysis, the primary model was refitted in the corresponding complete-case sample. β represents the interaction between race and WMH burden; 95% confidence intervals were calculated using heteroskedasticity-consistent (HC3) standard errors. Percentage change refers to the change in β magnitude relative to the corresponding primary-model estimate in the same complete-case sample. *P* values were adjusted for false discovery rate. The cohort-specific AD biomarker was CSF p-tau181/Aβ42 in EHBS and amyloid PET Centiloid in NACC. Vascular risk factors included hypertension, diabetes, dyslipidemia and obesity; arterial stiffness was assessed using pulse wave velocity. Adjustment for Area Deprivation Index was evaluated only in EHBS because coverage in NACC was limited and varied substantially by race and center. AD, Alzheimer’s disease; Aβ42, amyloid-β 1–42; *APOE*, apolipoprotein E; CSF, cerebrospinal fluid; EHBS, Emory Healthy Brain Study; NACC, National Alzheimer’s Coordinating Center; NHA, non-Hispanic African American; PET, positron emission tomography; p-tau181, tau phosphorylated at threonine 181; WMH, white matter hyperintensities.

## References

1. Duering, M. et al. Neuroimaging standards for research into small vessel disease—advances since 2013. The Lancet Neurology 22, 602–618 (2023).

2. Debette, S. & Markus, H. S. The clinical importance of white matter hyperintensities on brain magnetic resonance imaging: Systematic review and meta-analysis. BMJ 341, c3666–c3666 (2010).

3. Jansma, A., De Bresser, J., Schoones, J. W., Van Heemst, D. & Akintola, A. A. Sporadic cerebral small vessel disease and cognitive decline in healthy older adults: A systematic review and meta-analysis. Journal of Cerebral Blood Flow & Metabolism 44, 660–679 (2024).

4. Zahodne, L. et al. Structural MRI Predictors of Late-Life Cognition Differ Across African Americans, Hispanics, and Whites. Current Alzheimer Research 12, 632–639 (2015).

5. Lao, P. J. et al. Amyloid, cerebrovascular disease, and neurodegeneration biomarkers are associated with cognitive trajectories in a racially and ethnically diverse, community-based sample. Neurobiology of Aging 117, 83–96 (2022).

6. Stavitsky, K. et al. White Matter Hyperintensity and Cognitive Functioning in the Racial and Ethnic Minority Cohort of the Framingham Heart Study. Neuroepidemiology 35, 117–122 (2010).

7. Royse, S. K. et al. Unhealthy white matter connectivity, cognition, and racialization in older adults. Alzheimer’s & Dementia 20, 1483–1496 (2024).

8. Gavett, B. E. et al. Ethnoracial differences in brain structure change and cognitive change. Neuropsychology 32, 529–540 (2018).

9. Avila, J. F. et al. Education differentially contributes to cognitive reserve across racial/ethnic groups. Alzheimer’s & Dementia 17, 70–80 (2021).

10. Phuah, C.-L. et al. Association of Data-Driven White Matter Hyperintensity Spatial Signatures With Distinct Cerebral Small Vessel Disease Etiologies. Neurology 99, (2022).

11. Biesbroek, J. M. et al. Amyloid pathology and vascular risk are associated with distinct patterns of cerebral white matter hyperintensities: A multicenter study in 3132 memory clinic patients. Alzheimer’s & Dementia 20, 2980–2989 (2024).

12. Parent, O. et al. Characterizing spatiotemporal white matter hyperintensity pathophysiology in vivo to disentangle vascular and neurodegenerative contributions. Nature Communications 17, 4623 (2026).

13. Beyer, F. et al. White matter hyperintensity spatial patterns: Risk factors and clinical correlates. Alzheimer’s & Dementia 21, e70053 (2025).

14. Coenen, M. et al. Strategic white matter hyperintensity locations for cognitive impairment: A multicenter lesion-symptom mapping study in 3525 memory clinic patients. Alzheimer’s & Dementia 19, 2420–2432 (2023).

15. Rotblatt, L. J. et al. Impact of regional white matter hyperintensity patterns on cognitive trajectories in NACC. Alzheimer’s & Dementia 21, e70764 (2025).

16. Tremont, G., Halpert, S., Javorsky, D. J. & Stern, R. A. Differential Impact of Executive Dysfunction on Verbal List Learning and Story Recall. The Clinical Neuropsychologist 14, 295–302 (2000).

17. Wicklund, A. H., Johnson, N., Rademaker, A., Weitner, B. B. & Weintraub, S. Word List Versus Story Memory in Alzheimer Disease and Frontotemporal Dementia. Alzheimer Disease & Associated Disorders 20, 86–92 (2006).

18. Rizvi, B. et al. Association of Regional White Matter Hyperintensities With Longitudinal Alzheimer-Like Pattern of Neurodegeneration in Older Adults. JAMA Network Open 4, e2125166 (2021).

19. Jack, C. R. et al. Defining imaging biomarker cut points for brain aging and Alzheimer’s disease. Alzheimer’s & Dementia 13, 205–216 (2017).

20. Armstrong, M. K. et al. The association of structural versus load-dependent large artery stiffness mechanisms with cerebrovascular damage and cortical atrophy in humans. GeroScience 46, 5587–5597 (2024).

21. Hannawi, Y. et al. Association of Vascular Properties With the Brain White Matter Hyperintensity in Middle-Aged Population. Journal of the American Heart Association 11, e024606 (2022).

22. Towfighi, A. et al. Recommendations on Social Determinants of Health in Neurologic Disease. Neurology 101, (2023).

23. Walsemann, K. M., Jackson, H., Abbruzzi, E. & Ailshire, J. A. State-Level Education Quality and Trajectories of Cognitive Function by Race and Educational Attainment. The Milbank Quarterly 102, 765–821 (2024).

24. Goldstein, F. C., Hanfelt, J. J., James, T. A., Lah, J. J. & Loring, D. W. Receptive vocabulary is superior to education level to account for Black and White neuropsychological performance discrepancies. Journal of the International Neuropsychological Society 30, 998–1002 (2024).

25. Zahodne, L. B. et al. Longitudinal associations between racial discrimination and hippocampal and white matter hyperintensity volumes among older Black adults. Social Science & Medicine 316, 114789 (2023).

26. Zhu, Y. et al. Structural and social determinants of health shape the impact of neurodegeneration and cerebrovascular pathology on cognitive functioning in black adults. Social Science & Medicine 406, 119636 (2026).

27. Adkins-Jackson, P. B. et al. The structural and social determinants of Alzheimer’s disease related dementias. Alzheimer’s & Dementia 19, 3171–3185 (2023).

28. Hayes, C. A., Prasad, S., Dharmapuri, A., Thorpe, R. J. & The Health and Aging Brain Study (HABS-HD) Study Team. White matter hyperintensities mediate a greater proportion of age-related lower cognitive scores in non-Hispanic White participants compared to non-Hispanic Black and Hispanic participants: HABS-HD. *Alzheimer’s & Dementia: Diagnosis*, Assessment & Disease Monitoring 18, e70353 (2026).

29. Goetz, M. E. et al. Rationale and Design of the Emory Healthy Aging and Emory Healthy Brain Studies. Neuroepidemiology 53, 187–200 (2019).

30. Beekly, D. L. et al. The National Alzheimer’s Coordinating Center (NACC) Database: The Uniform Data Set. Alzheimer Disease & Associated Disorders 21, 249–258 (2007).

31. Besser, L. et al. Version 3 of the National Alzheimer’s Coordinating Center’s Uniform Data Set. Alzheimer Disease & Associated Disorders 32, 351–358 (2018).

32. Petersen, M. E. et al. Health and Aging Brain Study–Health Disparities (HABS-HD) methods and partner characteristics. Alzheimer’s & Dementia: Translational Research & Clinical Interventions 11, e70140 (2025).

33. Wu, J. et al. Benchmark White Matter Hyperintensity Segmentation Methods Fail on Heterogeneous Clinical MRI: A New Dataset and Deep Learning–Based Solutions. Journal of Imaging Informatics in Medicine https://doi.org/10.1007/s10278-025-01808-9 (2026) doi:10.1007/s10278-025-01808-9.

34. Isensee, F., Jaeger, P. F., Kohl, S. A. A., Petersen, J. & Maier-Hein, K. H. nnU-Net: A self-configuring method for deep learning-based biomedical image segmentation. Nature Methods 18, 203–211 (2021).

35. Fischl, B. FreeSurfer. NeuroImage 62, 774–781 (2012).

36. Sudre, C. H. et al. Bullseye’s representation of cerebral white matter hyperintensities. Journal of Neuroradiology 45, 114–122 (2018).

37. James, T. A. et al. Visual memory correlates with AD biomarkers in cognitively unimpaired individuals. Alzheimer’s & Dementia 22, e71417 (2026).

38. Weintraub, S. et al. Version 3 of the Alzheimer Disease Centers’ Neuropsychological Test Battery in the Uniform Data Set (UDS). Alzheimer Disease & Associated Disorders 32, 10–17 (2018).

39. Lee, M. L. et al. Bridging the gap: Addressing NACC’s evolving cognition batteries across UDS versions. Alzheimer’s & Dementia 21, e70832 (2025).

40. Klunk, W. E. et al. The Centiloid Project: Standardizing quantitative amyloid plaque estimation by PET. Alzheimer’s & Dementia 11, 1 (2015).

41. Athey, S., Tibshirani, J. & Wager, S. Generalized random forests. The Annals of Statistics 47, (2019).

42. Benjamini, Y. & Hochberg, Y. Controlling the False Discovery Rate: A Practical and Powerful Approach to Multiple Testing. Journal of the Royal Statistical Society Series B: Statistical Methodology 57, 289–300 (1995).

